# ExPRSweb - An Online Repository with Polygenic Risk Scores for Common Health-related Exposures

**DOI:** 10.1101/2022.01.13.22269176

**Authors:** Ying Ma, Snehal Patil, Xiang Zhou, Bhramar Mukherjee, Lars G. Fritsche

**Author notes:** Correspondence should be addressed to L.G.F. These authors jointly supervised this work (Bhramar Mukherjee, Lars G. Fritsche).

## Abstract

Complex traits are influenced by genetic risk factors, lifestyle, and environmental variables, so called exposures. Some exposures, e.g., smoking or lipid levels, have common genetic modifiers identified in genome-wide association studies. Since measurements are often unfeasible, Exposure Polygenic Risk Scores (ExPRSs) offer an alternative to study the influence of exposures on various phenotypes. Here, we collected publicly available summary statistics for 28 exposures and applied four common PRS methods to generate ExPRSs in two large biobanks, the Michigan Genomics Initiative and the UK Biobank. We established ExPRS for 27 exposures and demonstrated their applicability in phenome-wide association studies and as predictors for common chronic conditions. Especially, the addition of multiple ExPRSs showed, for several chronic conditions, an improvement compared prediction models that only included traditional, disease-focused PRSs. To facilitate follow-up studies, we share all ExPRS constructs and generated results via an online repository called ExPRSweb.

## Introduction

A central challenge in genetics is to understand inherited factors underlying complex traits and disorders. Substantial efforts in the past decade, especially genome-wide association studies (GWAS), have successfully uncovered genetic variants associated with a plethora of traits^1^. However, translating these to disease etiology or to predict outcomes is not straightforward. Most genetic risk variants have weak and sparse marginal effects, accounting for only a small fraction of the phenotypic variation, even for highly heritable traits^2–4^. Consequently, incorporating information across genetic variants is necessary for assessing the predisposition of complex traits.

The construction of polygenic risk score (PRS) is among the widely used approaches to translate genetic information into a disease risk^5,6^. A PRS is formed as a summation of an individual’s risk alleles, weighted by the effect sizes obtained from an external GWAS. PRS methods rely on the polygenicity of complex traits and vary in data input, model assumptions, validation procedures, and whether functional annotations or pleiotropic information is incorporated^7^.

In addition to genetic risk factors, lifestyle, and environmental variables, so called exposures, can impact disease risks. For example, high body mass index, smoking, blood lipid levels, and pre-existing type 2 diabetes (T2D) were recognized as prominent risk factors for cardiovascular disease^8^, respiratory diseases^9^, and cancers^10,11^. Given the relevance for these often modifiable risk factors for morbidity and mortality, exposure information is pivotal for precision prevention^10^. However, even common exposures are not always available, especially when using electronic health records (EHRs). Furthermore, data can be prone to measurement error, bias, and non-random missingness ^12,13^. Yet, some exposures have a heritable component identifiable through GWAS^14,15^ and thus offer the opportunity to construct Exposure PRSs (ExPRSs).

As genetic proxies at the individual level, ExPRSs have been used in many applications, e.g., risk prediction and stratification^16–18^, predicting exposures^19^, instruments for Mendelian randomization analyses, or phenome-wide association studies (PheWASs)^20–23^. Including ExPRSs to prediction models could improve disease diagnosis, screening, therapeutic interventions, and precision medicine approaches. PheWAS with ExPRSs may identify clinical phenotypes associated with a modifiable exposure and thereby highlight diseases whose onset might be influenced by early intervention or behavioral / lifestyle modification^20^. Noteworthy, ExPRSs capture the genetic predisposition of an exposure assigned at birth, but not the environmental influence, thus leaving a large proportion of the exposure’s variance unexplained. Still, the identification of associations between diseases and ExPRSs may help to tease apart the interplay of genetic and environmental pathways through which they influence disease risk.

The emerging utility of PRSs is evidenced via the accumulation of more than 1,000 PRS-related articles indexed in PubMed since 2009 ^24^ and spurred by significant advances in PRS methods^7^. Despite the rise in popularity, their transition into clinical settings is often limited by lack of transparency, compatibility, and reproducibility across cohorts. Therefore, a ExPRS resource that integrates adequate information for constructing, evaluating, and utilizing ExPRSs to accelerate ExPRS-related research is desirable and necessary. Recently, we established “Cancer PRSweb”, an interactive, online repository with cancer PRSs for 35 common cancer traits^20^. Building upon our previous work, we present ExPRSweb, a uniform analytic framework and an extension of PRSweb that specifically focusses on ExPRSs for 28 common exposures.

By using available exposure GWAS summary statistics and two large biobanks, the Michigan Genomics Initiative (MGI) and the UK Biobank (UKB), we generated ExPRS with four methods varying in complexity and modeling (i.e., linkage disequilibrium clumping and p-value thresholding [C+T], Lassosum, Deterministic Bayesian Sparse Linear Mixed Model [DBSLMM], and PRS-CS, a Bayesian method with continuous shrinkage priors)^25–30^. We also highlight ExPRSs applications including PheWAS, risk stratification and prediction of common chronic conditions. For the latter, we evaluated the predictive performance of single and multiple ExPRSs when combined with disease specific PRS and could show substantial improvement for several trait. We also contrasted these predictors with “Poly-Exposure Risk Scores” (PXSs) which integrate multiple measured exposures.

In absence of high-quality exposure data on many individuals ExPRSs can serve as surrogates if one has genotype data on a larger and more representative sample. Our repository ExPRSweb unlocks access to over 300 ExPRSs for 27 different exposures and facilitates scientific collaboration to strengthen their future application.

## Results

### Descriptive Characteristics of Study Cohorts

For the generation and analysis of ExPRSs, we used two analytical datasets that were restricted to unrelated participants of broad European ancestry encompassing 46,782 individuals in MGI and 408,595 individuals in UKB (**Table 1**; **Subjects and Methods**)^31–33^. The different prevalence of binary exposures and common chronic conditions in MGI and UKB likely reflect the characteristics of a hospital-based study (MGI) and a healthier, population-based study (UKB), respectively (**Table 1, Table S1**). For example, there are marked differences between MGI and UKB regarding hypertension (49.8% vs. 27.0%), diabetes (21.4% vs. 7.2%), and lung cancer (2.2% vs. 1.0%). Also, overweight individuals (74.7% vs. 66.8%) and smokers (49.2% vs. 39.4%) were more frequent in MGI (**Figure S1**).

**Table 1.**
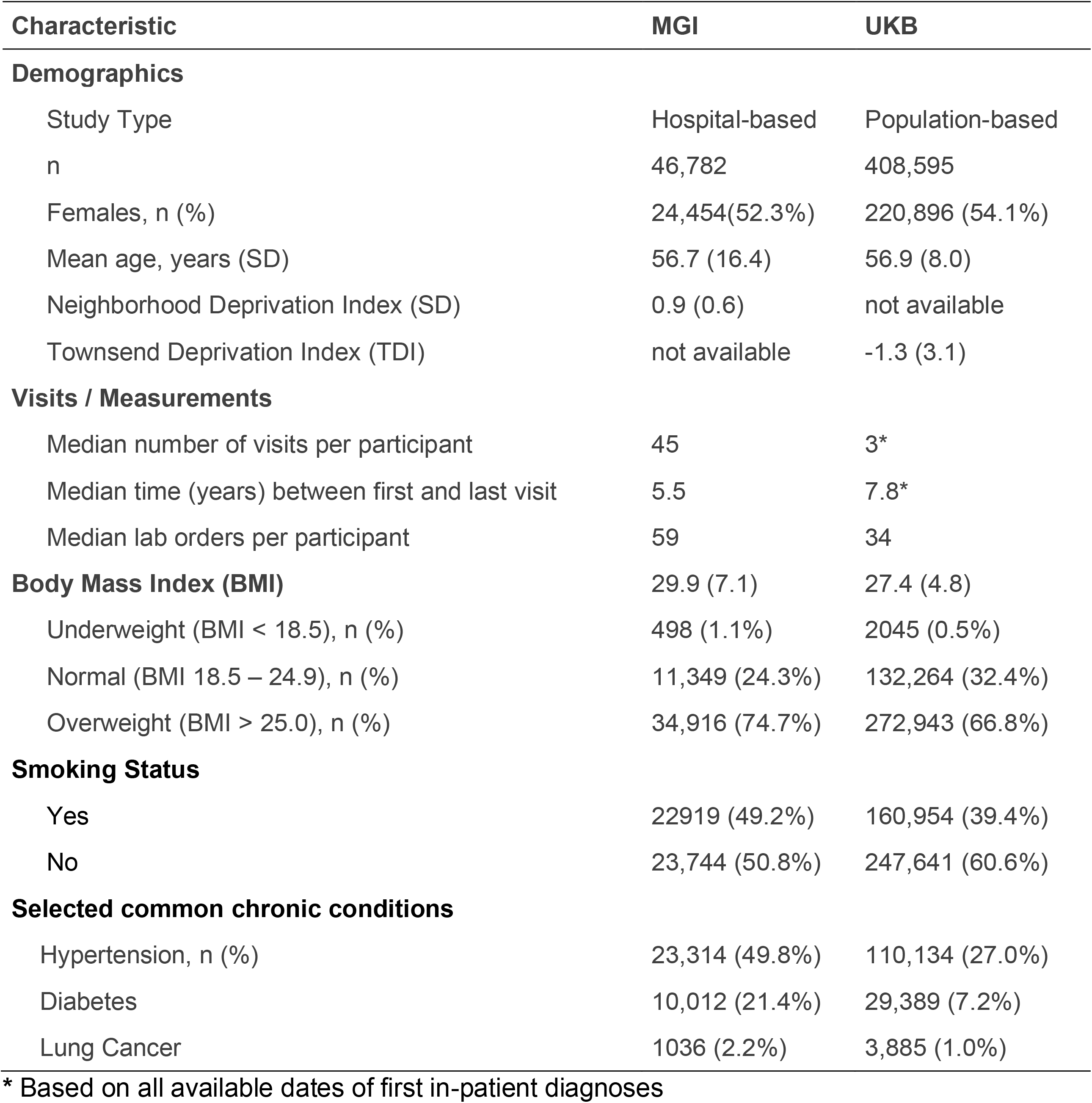
Demographics and Clinical Characteristics of the Analytic Datasets.

### Heritability Estimates

In total, we identified 82 sets of GWAS summary statistics for 28 different exposures (21 quantitative, 7 binary) that had matching exposure data in MGI and/or UKB; 52 solely based on UKB data and 30 on large GWAS (**Table 2, Table S2**). For each set, we estimated the narrow sense heritability^34^ as PRSs are closely connected to it and because one PRS method (DBSLMM) relies on these estimates. After excluding three GWAS sets with negative h^2^ estimates, we observed heritability estimates between 0.003 (sleep apnea) and 0.518 (height) that were in line with previous studies (**Table S3**) ^4,35–39^. Still, estimates from GWAS on the same exposure often varied (e.g., h^2^[height]: 0.012 – 0.518 or h^2^[vitamin D]: 0.009 – 0.100) implying different underlying frameworks (**Figure S2)**.

**Table 2.**
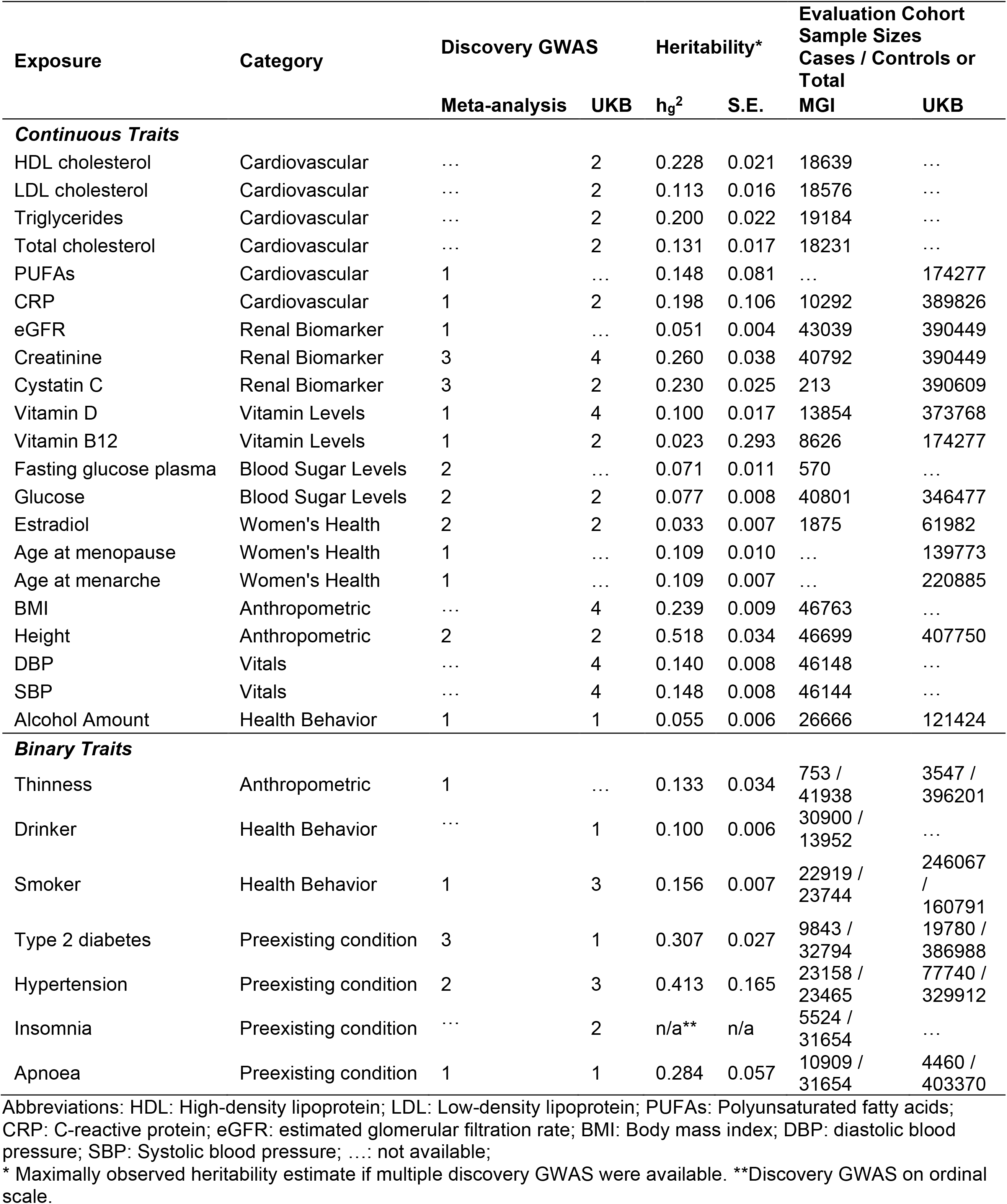
Overview of the 28 Included Exposures Traits. Number of included discovery GWASs, estimated heritability (liability scale) and sample size of PRS evaluation cohorts are shown. Details can be found in Tables S1-S3.

### ExPRS Evaluation

Following the scheme in **Figure 1**, we generated 514 ExPRSs (379 for 25 exposures in MGI and 135 for 17 exposures in UKB; **Table S4**) and assessed association, overall performance, accuracy, and discrimination. A total of 336 ExPRSs for 27 exposures were nominally significant and positively associated with their corresponding exposures in MGI (262 ExPRSs; 24 exposures) and in UKB (74 ExPRSs; 14 exposures) and analyzed further (**Table S4**).

**Figure 1.**
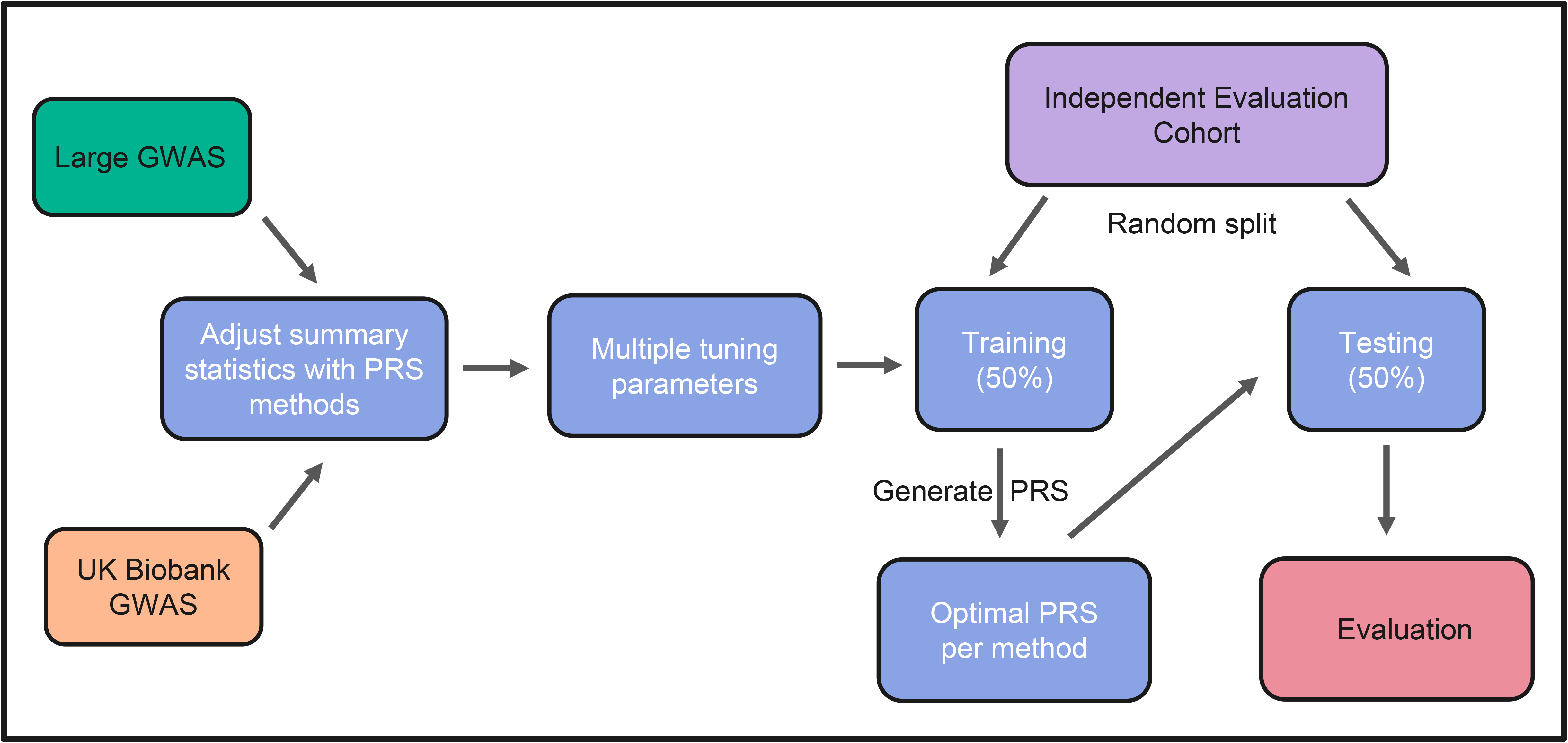
Flow chart of ExPRS construction, evaluation, and selection.

#### Performance comparison across methods

For the method comparison we focus on MGI having a more comprehensive set of exposures covered by ExPRSs. PRS-CS produced the best performing ExPRSs for 18 of the 24 exposures consistent with previous benchmarking (**Table 3**, **Figure 2, and Figure S3**)^40–42^. Lassosum excelled for the alcohol and smoker exposure, DBSLMM for lipid levels, and both C+T implementations for exposures with low heritability, e.g., vitamin B12 and D (**Figure 2**). Further, we found that the C+T implementation that uses dosages for LD clumping had a slight edge over the one using best guess genotypes, confirming previous findings^43^. Overall, these results suggested the methods’ performances differed by trait showcasing the benefit of screening multiple methods.

**Figure 2.**
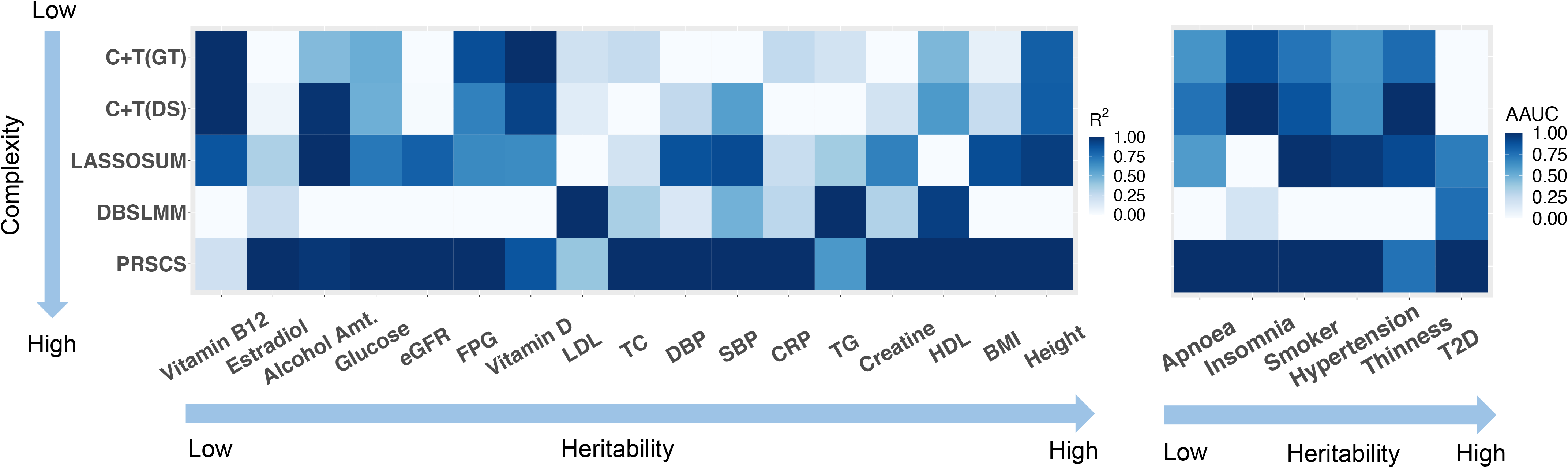
Prediction performance of the five applied PRS methods in MGI across continuous (left) and binary (right) traits. Here, the heatmap shows the relative prediction performance for each method across traits (values were scaled to 0-1 range) for better comparison. Specifically, the prediction performance is quantified using *R*^2^ for continuous traits and covariate-adjusted AUC for binary traits. For a fair comparison we selected the same summary statistic for each method (GWAS with the highest heritability estimate).

**Table 3.** Top ranked ExPRS in MGI. Details about the underlying discovery GWAS study, can be found in Table S2.

| Exposure | Discovery GWAS | Method | # SNPs in ExPRS | Association P | Brier Score | Pearson's r | Adjusted AUC (95% CI) |
| --- | --- | --- | --- | --- | --- | --- | --- |
| **Continuous Traits** |  |  |  |  |  |  |  |
| HDL | UK Biobank | PRS-CS | 1113830 | 1.3E-294 | ... | 0.311 | ... |
| LDL | UK Biobank | DBSLMM | 8918470 | 4.1E-174 | ... | 0.274 | ... |
| TG | UK Biobank | DBSLMM | 8924773 | 6.9E-304 | ... | 0.348 | ... |
| TC | UK Biobank | PRS-CS | 1113831 | 3.2E-168 | ... | 0.265 | ... |
| CRP | UK Biobank | PRS-CS | 1113831 | 6.2E-30 | ... | 0.155 | ... |
| eGFR | Meta-analysis | PRS-CS | 1113831 | 3.8E-150 | ... | 0.13 | ... |
| Creatinine | UK Biobank | PRS-CS | 1113831 | 4.6E-189 | ... | 0.174 | ... |
| Vitamin D | UK Biobank | C+T(GT) | 500 | 9.0E-46 | ... | 0.166 | ... |
| Vitamin B12 | UK Biobank | C+T(DS) | 9 | 0.0011 | ... | 0.049 | ... |
| FPG | Meta-analysis | C+T(GT) | 273 | 0.0095 | ... | 0.099 | ... |
| Glucose | UK Biobank | PRS-CS | 1113830 | 8.4E-112 | ... | 0.156 | ... |
| Estradiol | UK Biobank | PRS-CS | 1113823 | 0.046 | ... | 0.0664 | ... |
| BMI | UK Biobank | PRS-CS | 1113832 | 5.3E-607 | ... | 0.319 | ... |
| Height | UK Biobank | PRS-CS | 1113832 | 2.2E-1988 | ... | 0.373 | ... |
| DBP | UK Biobank | PRS-CS | 1113831 | 4.8E-170 | ... | 0.166 | ... |
| SBP | UK Biobank | PRS-CS | 1113831 | 1.7E-189 | ... | 0.172 | ... |
| Alcohol Amt. | Meta-analysis | PRS-CS | 1116497 | 1.4E-18 | ... | 0.0746 | ... |
| **Binary Traits** |  |  |  |  |  |  |  |
| Thinness | Meta-analysis | C+T(DS) | 256 | 0.044 | 0.018 | ... | 0.532<br>(0.503, 0.561) |
| Drinker | UK Biobank | PRS-CS | 1113832 | 5.1E-29 | 0.212 | ... | 0.547<br>(0.539, 0.536) |
| Smoker | Meta-analysis | PRS-CS | 1109786 | 1.10E-170 | 0.232 | ... | 0.605<br>(0.598, 0.612) |
| T2D | Meta-analysis | PRS-CS | 945820 | 1.10E-159 | 0.139 | ... | 0.637<br>(0.621, 0.653) |
| Hypertension | UK Biobank | PRS-CS | 1113832 | 6.4E-213 | 0.182 | ... | 0.630<br>(0.622, 0.639) |
| Insomnia | UK Biobank | PRS-CS | 1065129 | 5.0E-06 | 0.126 | ... | 0.524<br>(0.513, 0.536) |
| Sleep apnea | UK Biobank | PRS-CS | 1111194 | 5.8E-10 | 0.182 | ... | 0.527<br>(0.517, 0.536) |

Abbreviations: HDL: High-density lipoprotein; LDL: Low-density lipoprotein; PUFAs: Polyunsaturated fatty acids; CRP: C-reactive protein; eGFR: estimated glomerular filtration rate; BMI: Body mass index; DBP: diastolic blood pressure; SBP: Systolic blood pressure; ...: not applicable.

#### Performance across exposures

Again, focusing on MGI, we selected for each exposure the ExPRS with the lowest association p-value among its method / exposure GWAS combinations (**Table S4**). For quantitative exposures the Pearson’s correlation r with their corresponding ExPRS ranged from 0.049 (Vitamin B12) to 0.373 (Height). For binary exposures, the area under the covariate-adjusted area under the ROC curve (AAUC) ranged from 0.524 (insomnia) to 0.637 (T2D), confirming only modest discrimination by PRS for complex traits ^44^. The ExPRSs’ performance generally agreed with the ranking of their heritability estimates (**Figure S4** and **S5**).

#### Performance comparison across cohorts

Like MGI, we selected in UKB for each of the 14 exposures the ExPRS that reached the strongest association (**Table S5**): 6 were based on Lassosum, 4 on PRS-CS, 3 on C+T (DS), and 1 on C+T (GT). In contrast to MGI, Lassosum outperformed the other methods in UKB (**Figure S6** and **S7**). The inconsistencies across cohorts might be the result of different underlying GWAS sets, i.e., for UKB ExPRSs we only relied on non-UKB studies to avoid overfitting. Also, the methods’ varying tuning procedures especially for Lassosum and C+T might be affected by the larger sample sizes in UKB. For ExPRSs of quantitative exposures, the correlation with their corresponding exposures ranged from 0.015 (alcohol consumption) to 0.326 (height) (**Table S5**). For binary exposures, the AAUC ranged from 0.505 (hypertension) to 0.825 (T2D). When comparing ExPRSs on exposures that were present in both cohorts, we found generally consistent performances for quantitative traits like C-reactive protein, creatine, vitamin D, and height, while for some binary traits such as the T2D (AAUC_MGI_: 0.64, AAUC_UKB_: 0.83) and smoking (AAUC_MGI_: 0.61, AAUC_UKB_: 0.77), AAUC differed substantially (**Table S6**). Noteworthy, the estimates in UKB might be heightened due to undetected, overlapping samples between their discovery GWAS and the UKB cohort^45,46^, or caused by to the cohort’s larger effective sample sizes.

### Correlations of ExPRSs across exposures

Next, we assessed the relationships between ExPRSs and exposures in MGI. **Figure 3** displays the pairwise correlation between 15 quantitative exposures, between their 15 corresponding ExPRSs, and between the ExPRSs and the quantitative exposures in MGI. The correlations between the quantitative exposures indicated positive and negative relationships (r between -0.1 and 0.92; **Figure 3A**), the strongest between closely related exposures: r[TC, LDL] = 0.92, r[eGFR, creatine] = -0.84, and r[SBP, DBP] = 0.53. The former two can be attributed to their underlying equations and related measurements, while the linear relationship between SBP and DBP is well-established^47–49^. Several of the other observed correlations are also well-documented often reflecting related disease etiologies ^50–52^. Similar but more attenuated patterns were seen for the ExPRSs whose correlations ranged from -0.78 to 0.72 (**Figure 3B**). The often lower pairwise correlations (e.g., r[PRS_TC_, PRS_LDL_] = 0.72 and r[PRS_eGFR_, PRS_creatine_] = -0.78) were expected because ExPRSs capture only a fraction of the exposure’s variance (see diagonal of **Figure 3C**). The consistent patterns suggested that several ExPRSs can replicate correlations of measured exposures relatively well and thus might be suitable surrogates for exposures, especially for studies where measurements might not be feasible or likely be biased ^50,53,54^.

**Figure 3.**
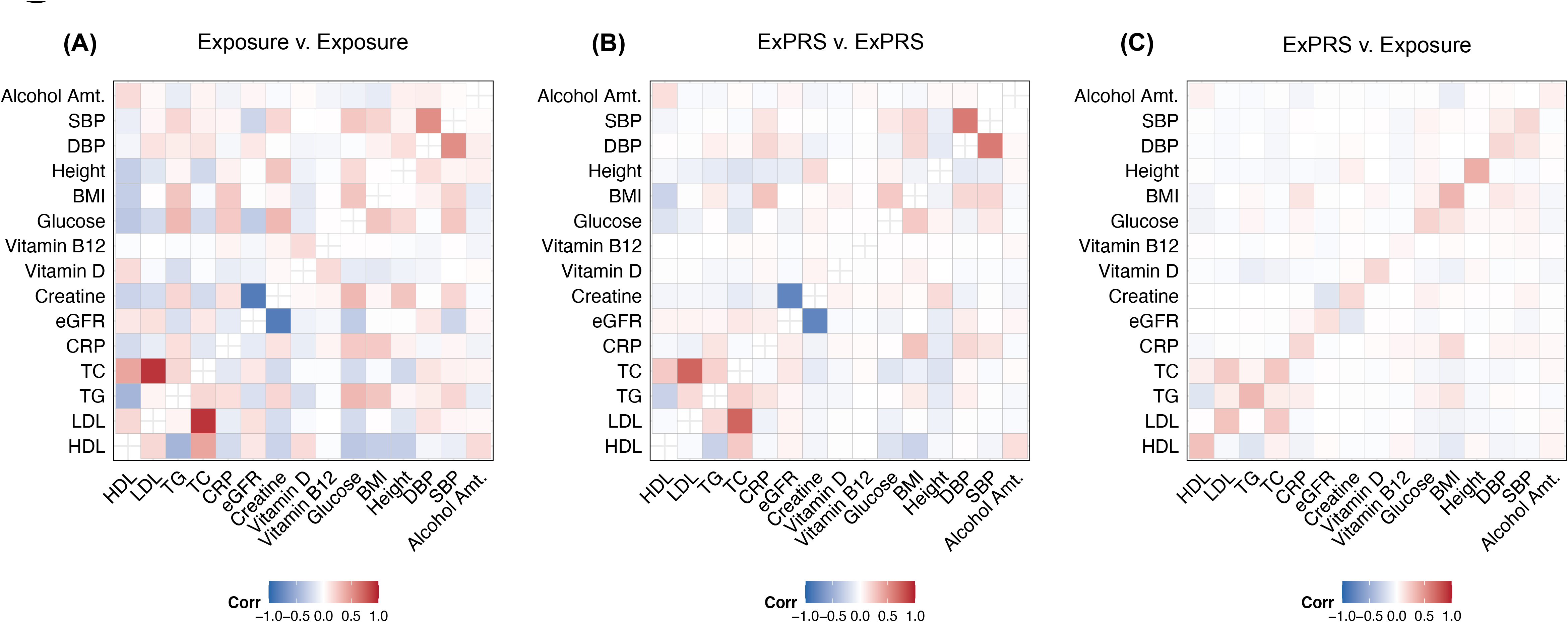
Comparison of the pairwise correlation of 15 ExPRS and their corresponding continuous traits in MGI. Heatmap displays the pairwise correlation between (A)15 continuous exposures in MGI; (B) ExPRSs; and (C) Exposures (y-axis) and ExPRSs (x-axis). Here, Pairwise Spearman correlation with nominally significant association P values (<= 0.05) are shown. Fasting plasma glucose (exposure and ExPRS) was excluded due to the exposures low sample size in MGI.

### ExPRS Applications

#### Phenome-wide Association Analyses

One application of ExPRSs is their use as predictors for phenome-wide association studies (PheWASs) to uncover phenotypes with a shared genetic component and thus disorders that might benefit from an early intervention. We showcase such ExPRS PheWASs by analyzing all 24 selected ExPRSs across up to 1685 EHR-derived phenotypes (PheCodes) in MGI (**Table S7**). In total, we observed phenome-wide significant associations between 22 ExPRSs and 440 phenotypes (Bonferroni-corrected threshold at P < 0.05 / 1685; **Table S8**). Overall, the number and the strength of observed associations seem to depend on the exposures’ impact and heritability. For example, the PheWAS with the BMI ExPRS uncovered 329 associated phenotypes while the Vitamin B-12 ExPRS PheWAS only revealed two associations with closely related phenotypes. Besides the expected associations between BMI PRS and obesity-related phenotypes (1.66 < OR < 2.14, e.g., obesity, morbid obesity, and overweight), we also observed significant phenome-wide associations with hypertension (OR: 1.33 [1.30,1.36]), T2D (OR: 1.41 [1.37,1.45]), osteoarthrosis (OR: 1.15 [1.12,1.17]), and sleep apnea (OR: 1.28 [1.25,1.31]); all previously reported for BMI^55–58^ **(Figure 4A; Table S8**). The PheWAS with measured BMI revealed consistent associations (**Figure 4C; Table S9**) though with larger effects: hypertension (OR: 1.88 [1.84, 1.93]), type 2 diabetes (OR: 2.00 [1.95, 2.06]), osteoarthrosis (OR: 1.29 [1.27,1.32]), and sleep apnea (OR: 2.24 [2.18, 2.30]).

**Figure 4.**
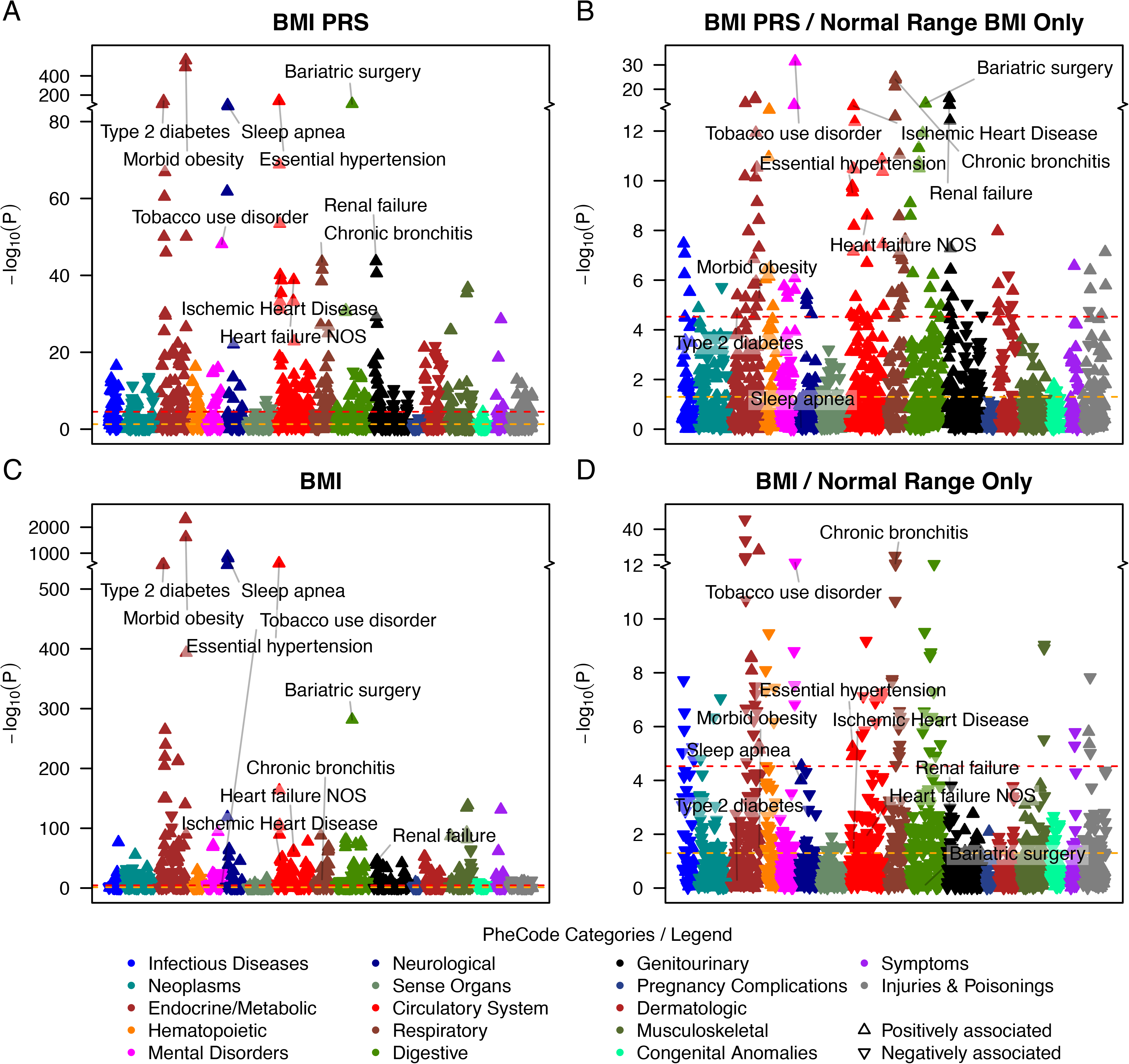
ExPRS PheWAS and Exclusion ExPRS PheWAS as an example for continuous traits in MGI. (A) ExPRS PheWAS plot for BMI ExPRS (B) Exclusion ExPRS PheWAS plot is shown for using BMI ExPRS as predictor among the individuals with normal BMI value (18.5 ∼ 24.9 kg/ m2) (C) Trait PheWAS plot is shown for BMI trait (D) Exclusion Trait PheWAS plot is shown for using BMI trait as predictor among the individuals with normal BMI value. The axis breaks were chosen so that the 10 strongest signals fall in the top scale (y-axis breaks for the four panels at -log_10_P are 84, 13, 540, and 12, respectively).

To assess whether these associations were driven by exposed individuals, i.e., individuals affected by a binary exposure or by low or high exposure values, we also performed “Exclusion-PRS-PheWAS” analyses where we excluded such exposed individuals to remove direct and indirect associations of the exposure and potential treatment effects (see **Methods**). While this exclusion of individuals markedly decreased sample sizes and thus power, we identified 198 phenotypes that remained significantly associated with 17 ExPRSs in the Exclusion PRS PheWAS (P < 0.05 / 1685; **Table S8**). For example, in the Exclusion PheWAS with the BMI ExPRS the associations with hypertension (OR: 1.17 [1.12, 1.23]) and T2D (OR: 1.18 [1.09, 1.27]) remained statistically significant (**Figure 4B, Table S8**). However, while the analysis of individuals with healthy BMI removed most of the obesity or overweight phenotypes, a strong association remained between BMI ExPRS and bariatric surgery (OR: 2.66 [2.08, 3.41]). A closer inspection revealed that 73 of 1509 MGI participants who underwent bariatric surgery had recorded median BMI values that fell in the healthy BMI range (18.5 < BMI < 25) indicating the BMI ExPRS’s ability to capture pre-treatment exposures. Most interestingly, the corresponding Exclusion PheWAS with measured BMI as predictor revealed many association signals that were reversed compared to the Exclusion-PRS-PheWAS (**Figure 4C** and **4D**, **Table S8** and **S9)**. This finding might reflect biased measurements, e.g., due to treatment or interventions that result in normal BMI values, or the measured BMI’s inability to capture central obesity^59^.

We performed similar sets of PheWASs in UKB. While based on a separate ExPRS generations restricted to UKB-independent GWAS summary statistics, most of the strong associations seen in the MGI were also seen in the UKB ExPRS PheWASs, e.g., obesity associated with T2D PRS (OR_MGI_: 1.71 [1.15, 1.20] and OR_UKB_: 1.63 [1.60, 1.66]; **Figure S8** and **S9**). Due to the larger sample sizes in the UKB compared to MGI (**Table 2**), we often observed more and stronger secondary trait associations (**Table S10** and **S11**).

In general, we found that agnostic ExPRS PheWASs can provide valuable insights into exposure – phenotype relationships, many of which were previously reported for measured exposures. However, thorough investigations are needed to distinguish between spurious and genuine signals.

#### Improving Prediction Models for Common Chronic Conditions

As many exposures are important risk factors for common chronic conditions^9,60,61^, we performed analyses with a specific emphasis on 27 chronic conditions whose algorithms are used for Medicaid and Medicare claims and available from the Chronic Condition Data Warehouse (CCW, **Table S12**) ^62^. Since these were developed for the US health system and lack transferability to the UK, we limited our analysis to MGI. Related chronic conditions were already covered in the phenome-wide PheCode-based association analyses, therefore this targeted analysis of “real world” phenotype algorithms aims to evaluate the ExPRSs’ abilities to improve predictions. Basically, we are interested to see if a prediction model that solely relies on a GWAS-based PRS for a chronic condition “Y” (YPRS) can be augmented with additional ExPRSs.

As a first step, we explored the association between the 27 conditions and the 24 ExPRSs. We found that even after excluding the directly related condition/exposure pairs (e.g., hypertension/SBP ExPRS, Hyperlipidemia/TC ExPRS, etc.) all included 24 ExPRSs showed a nominally significant association with at least one condition at P < 0.05 (**Table S13**). Vice versa, 26 of the 27 conditions were nominally significant associated with at least one ExPRS substantiating the exposures’ relevance. However, none of the ExPRSs was associated with Alzheimer’s Disease though many of the included exposures were reported risk factors ^63^. The strongest risk increasing effect was seen for BMI ExPRS and diabetes (OR: 1.393 [1.357, 1.430]), while the strongest protective effect was seen for HDL ExPRS and diabetes (OR: 0.823 [0.803, 0.844]) (**Table S13**).

Considering the relatively poor predictive performance of single ExPRSs for chronic conditions and that some of the chronic conditions were associated with several ExPRSs (**Table S13**), we next assessed whether the combination of ExPRS (“multiExPRS”, see **Methods**) can improve risk prediction of models that only include YPRSs (**Table S14**).

Due to the required cross-validation, limited sample sizes, and limited availability of YPRS (**Table S15**), we restricted our comparisons to 12 conditions (**Table S12** and **S15**). We found that adding multiple ExPRSs enhanced models for several conditions (e.g., stroke/transient ischemic attack, heart failure, lung cancer, hypertension, chronic kidney disease, asthma; **Table S16**, **Figure 5**). For example, the AAUC for predicting hypertension increased from 0.627 to 0.637 when adding multiple ExPRSs (BMI, C reactive protein, drinking status, fast plasma glucose, HDL, height, smoking status, T2D, triglycerides, apnea, and insomnia). In contrast, the addition of ExPRSs did not improve prediction accuracy for other conditions (e.g., glaucoma, prostate cancer, colorectal cancer, and atrial fibrillation). Nevertheless, the ability of specific ExPRSs to improve predictions indicate that some of the YPRS often do not capture the entirety of an individual’s genetic predisposition, likely reflecting the lack of power of the condition’s discovery GWAS compared to exposure GWASs which due to larger sample sizes and continuous measurements are often better powered.

**Figure 5.**
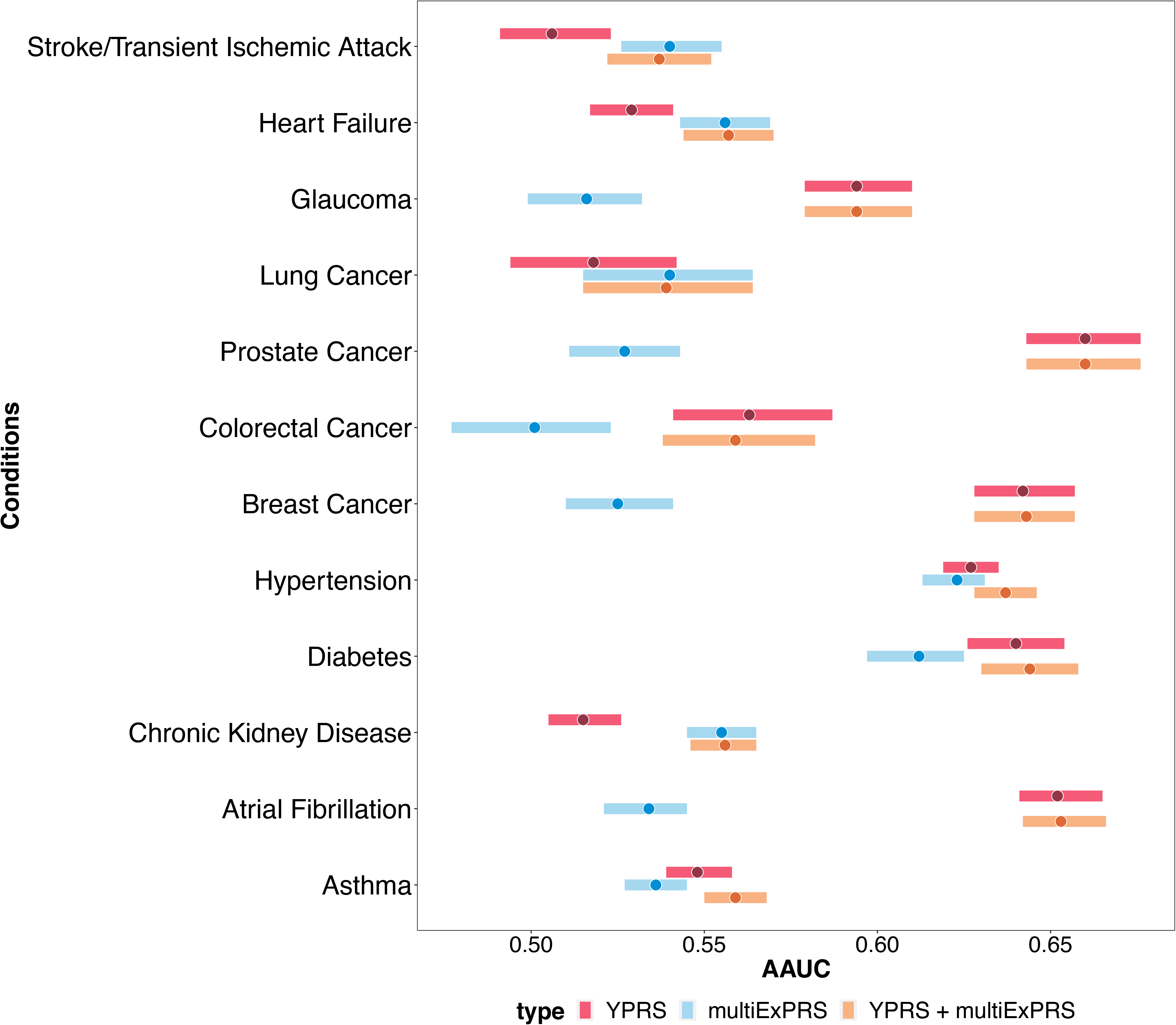
Comparisons of the prediction performance of different predictors for common chronic conditions in MGI cohort. AAUC paired with 95% confidence interval for condition specific PRS (YPRS, red), combined ExPRSs (multiExPRS, blue) and YPRS + multiExPRS (orange) were shown as forest plot. Each bar represents the 95% interval for the AAUC with the dot represents the AAUC estimate.

Since these predictions yielded only moderate to poor discrimination (AAUC < 0.66), we also evaluated the ExPRSs’ ability to augment risk stratification with YPRS, i.e., to define subsets of individuals at high risk for the 12 conditions (**Figure S10**, **Table S18** and **S17**). All YPRS were by themselves able to significantly enrich cases in their distributions’ tails, though not across all five top percentiles (1, 5, 10, and 25%). For example, only 8 YPRSs could significantly enrich cases in the top 1% at P < 0.05 with OR ranging from 1.79 (95% CI: 1.37, 2.35; asthma) to 7.13 (95% CI: 5.57, 9.13; atrial fibrillation).

Adding the combined ExPRSs (multiExPRSs) to the “YPRS only model” improved the enrichment of cases for 10 of the 12 conditions when considering the top 1%. A major improvement was seen for the enrichment of cases with heart failure (YPRS: OR: 1.13 [0.74, 1.72] vs YPRS + multiExPRS: OR: 2.72 [1.94,3.81]) and with hypertension (YPRS: OR: 2.96 [2.31,3.80] YPRS + multiExPRS: OR: 3.73 [2.94,4.73]). However, adding multiple ExPRS negatively affected the enrichment of cases with atrial fibrillation (YPRS: OR: 7.13 [5.57, 9.13] vs YPRS + multiExPRS: OR: 6.21 [4.82, 8.00]). Similar but less pronounced enrichments of cases were seen for the top 2%, top 5%, top 10%, and top 25% (**Figure S10**, **Table S18**).

Our explorations confirmed that individuals in the tails of PRS distributions are most informative for risks of chronic conditions^64^. Further, the consistent gain in risk stratification by adding multiple ExPRS highlights their potential use.

Finally, we compared the application of the PRSs (YPRs and/or the multiExPRSs) with poly-exposure scores (PXSs) that are based on measured / collected exposure data as previously described for type 2 diabetes ^65^. Again, focusing on the 12 conditions (**Table S12**), we created a PXS for each condition in the MGI cohort using up to 24 of 27 available exposures (**Subjects and Methods**). The number of incorporated exposures ranged from 7 (glaucoma) to 19 (chronic kidney disease) (**Table S19** and **S20**). Though the evaluation cohorts were different in size, we observed that the PXSs mostly showed better discrimination than the models that only relied on PRSs (YPRS, multiExPRS, or YPRS + multiExPRS), except for colorectal, prostate, and female breast cancer which underperformed (**Figure S11, Table S18**). Since PXSs were only obtainable for people who had complete data for each included exposure, they were only available for a small fraction of the genotyped MGI individuals for which YPRSs and ExPRSs were obtainable (2.5 - 18.4%; Glaucoma: 56.0%). Furthermore, the proportion of genotyped individuals with complete exposure data for their PXSs was significantly different between cases and controls for 9 of the 12 analyzed conditions, indicating non-random missingness of exposures in the MGI EHR that likely biased the analysis. The most extreme example was chronic kidney disease with cases being about four times more likely than controls to have complete exposure data for their PXS (OR 3.9 [3.5, 4.4], P = 31.x10^-107^; **Table S21**).

### Online Visual Catalog: *ExPRSweb*

In our current study, we generated and evaluated hundreds of ExPRSs in which predictive properties differed between GWAS source, exposure, method, and/or evaluation cohort (**Table S4**). To enable an exploration of the ExPRS for 27 different exposures, we created a new PRSweb ^20^ instance called ExPRSweb (see **Web Resources**) that includes detailed metrics (association, performance, discrimination, and accuracy) and allows the selection of ExPRSs based on properties for specific applications. The tables, like **Table 3** and **Table S5**, can be sorted, filtered, or downloaded. ExPRSweb also offers detailed information about each ExPRS, including GWAS source(s), LD reference panels and the included risk variants, effect/non-effect alleles, and weights. ExPRSweb also links to interactive ExPRS-PheWAS results for their evaluation cohort,

## Discussion

In this study, we have constructed and evaluated a large set of ExPRSs using 79 sets of GWAS summary statistics, applied various PRS methods and while doing so, created over 514 ExPRSs of which 336 showed promising performance for 27 different exposures in MGI and/or UKB.

We explored the performance of ExPRSs across methods, GWAS sources, and two cohorts and observed two key points that might be helpful to strategize future ExPRS generation projects: First, large exposure GWAS with higher SNP heritability estimates usually also resulted in the most predictive ExPRS. Second, there was no “one size fits all” recipe for ExPRSs, i.e., we recommend the screening of multiple methods.

While there is a wide range of health-related exposures ^66–68^, we focused on 28 exposures for which we could find GWAS from external full summary statistics and which sufficiently measured samples in MGI and/or UKB. The exposures can roughly be categorized into cardiovascular, renal biomarkers, vitamin levels, blood sugar levels, women’s health, anthropometric measurements, vitals, health behaviors, and preexisting conditions. However, other relevant exposure were not explored in this study, e.g., dietary exposures (e.g., milk consumption, coffee consumption^69,70^), telomere length^71^, and other biomarkers (e.g., Transforming growth factor beta (TGF-beta)^72,73^, circulating microRNA miR-34b (Ref.^74^). While some exposures also have GWAS summary statistics available (e.g., coffee consumption, milk consumption summary statistics from UKB GWAS efforts), the exposures were not measured in MGI and thus could not be evaluated at this time.

We generated ExPRSs using four methods (C+T, Lassosum, DBSLMM and PRS-CS), all of which are computationally efficient, but skipped other new methods that have been proposed (SBayesR, LDPred, NPS, SCT) ^75–78^; but often require massive computational resources, especially for large cohorts like UKB and MGI. Additionally, several alternative methods were reported to improve predictive power by incorporating external information (e.g., functional annotations, pleiotropy across multiple traits), e.g., LDpred-funct^79^, AnnoPred^80^ and MTGBLUP^81^. Future implementations and systematic evaluations of these alternative choices are needed to further the availability of well-powered ExPRSs and their applications.

We evaluated ExPRSs in cohorts of broadly European ancestry because of the limited diversity in MGI and UKB. However, the lacking transferability across ancestry groups increases the need to also construct ExPRS for non-European ancestry groups^29,77, 82–84^. While efforts are underway to develop cross-ancestry PRS methods to increase transferability, ultimately an increased diversity in datasets is needed to counteract the European ancestry bias in GWAS that is passed on to PRS research^85,86^.

Our explorations of ExPRSs, mainly in the MGI cohort, revealed that some of the ExPRSs could be good surrogates for exposures and enable meaningful association analyses across medical phenomes or a collection of chronic conditions. Also, the combination of ExPRSs can improve predictions and risk stratification beyond the YPRSs, e.g., for asthma, heart failure, or hypertension. Yet, for some of the studied conditions, the additional of multiple ExPRSs did not improve models that already included YPRSs. This suggests that YPRSs, if based on very large sample sizes, might already have captured most of the genetic risk profiles reflecting direct and indirect (exposure mediated) risk effects.

There are other applications of ExPRSs that we did not explore but that gained attention in the recent years, e.g., mediations analyses to study polygenic pleiotropy ^87^ or their use as instrumental variables in Mendelian Randomization analysis to uncover novel mechanisms that contribute towards disease susceptibility^88–91^.

A main application for ExPRSs might be their use as proxies for unmeasured exposures. Exposures relevant for many conditions are often only sparsely measured in the EHR data sets and their missingness can substantially reduce sample size when considering only complete case datasets (as seen here for PXS). Furthermore, contrary to genotype data the missingness can be non-random because testing generally is selective, diagnosis- and symptom-specific, as seen here for 9 of the 12 analyzed conditions, and thus likely would bias prediction models. Nevertheless, an ExPRS can even in the best scenario only capture the heritable fraction of the exposure’s variance coming from variants assigned at birth but not the lifelong exposure to environmental or consequences of behavioral factors. Using ExPRSs for the imputation of incomplete exposure data could be worth further explorations but was not within the scope of the current study.

Being dependent on large GWAS and evaluation cohorts we expect that future studies will provide more powerful YPRS and ExPRSs. But even then, the interplay of genetic and non-genetic factors needs to be considered when assessing complex traits. Current large biobank efforts link genotype data with EHRs, and often complement patient information on environmental, lifestyle, and demographic variables via self-report ^92^. The integration of these resources will likely improve our models with the goal to prevent or treat conditions earlier.

Finally, we created an online repository called “ExPRSweb” that similar to our cancer specific PRS repository “Cancer PRSweb” ^20^, provides an interactive platform to browse performance metrics of all generated ExPRSs in two independent biobanks. We also deposited all promising ExPRSs to the PGS catalog and linked it to ExPRSweb and our evaluations. We anticipate that ExPRSweb can serve as an example and a standardized platform to expedite ExPRS research and to facilitate easier access.

## Methods

### Michigan Genomics Initiative (MGI)

#### MGI Cohort

Adult participants aged between 18 and 101 years at enrollment were recruited through the Michigan Medicine health system between 2012 and 2020. Participants have consented to allow research on both their biospecimens and EHR data, as well as linking their EHR data to national data sources such as medical and pharmaceutical claims data. Participants were primarily recruited through the MGI - Anesthesiology Collection Effort while awaiting a diagnostic or interventional procedure either at a preoperative appointment or on the day of their operative procedure at Michigan Medicine. Additional participants were recruited through the Michigan Predictive Activity and Clinical Trajectories (MIPACT) Study, the Mental Health BioBank (MHB2), and the Michigan Genomics Initiative-Metabolism, Endocrinology, and Diabetes (MGI-MEND) Study. The data used in this study included diagnoses coded with the Ninth and Tenth Revision of the International Statistical Classification of Diseases (ICD9 and ICD10) with clinical modifications (ICD9-CM and ICD10-CM), laboratory measurements, anthropometrics (height, thinness and body mass index [BMI]), vitals (systolic and diastolic blood pressure [SBP and DBP]), health behavior (alcohol amount, smoker and drinker), sex, precomputed principal components (PCs), genotyping batch, recruitment study, and age. Data were collected according to the Declaration of Helsinki principles. MGI study participants’ consent forms and protocols were reviewed and approved by the University of Michigan Medical School Institutional Review Board (IRB ID HUM00099605 and HUM00155849). Opt-in written informed consent was obtained. Additional details about MGI can be found online (see **Web Resources**).

#### MGI Genotype Data

DNA from 56,984 blood samples was genotyped on customized Illumina Infinium CoreExome-24 bead arrays and subjected to various quality control filters, resulting in a set of 502,255 polymorphic variants. Principal components and ancestry were estimated by projecting all genotyped samples into the space of the principal components of the Human Genome Diversity Project reference panel using PLINK (v1.90b5.4, 938 individuals) ^93,94^. Pairwise kinship was assessed with the software KING (2.2.4) ^95^, and the software FastIndep was used to reduce the data to a maximal subset that contained no pairs of individuals with 3rd-or closer degree relationship ^96^. We removed participants without EHR data and participants not of recent European descent from the analysis, resulting in a final sample of 46,782 unrelated subjects. Additional genotypes were obtained using the Haplotype Reference Consortium reference panel of the Michigan Imputation Server ^97^ and included over 24 million imputed variants with R^2^ ≥0.3 and minor allele frequency (MAF) ≥0.01%.

#### MGI Phenome

The MGI phenome was based on ICD9-CM and ICD10-CM code data for 46,782 unrelated, genotyped individuals of recent European ancestry. Longitudinal time- stamped diagnoses were recoded to indicators for whether a patient ever had given a diagnosis code recorded by Michigan Medicine. These ICD9-CM and ICD10-CM codes were aggregated to form up to 1,814 PheCodes using the PheWAS R package (as described in detail elsewhere ^20,98,99^). To minimize differences in age and sex distributions, avoid extreme case-control ratios, and reduce the computational burden, we matched up to 10 controls to each case using the R package “MatchIt” ^100^. Nearest neighbor matching was applied for age and the first four principal components of the genotype data (PC1-4) using Mahalanobis distance with a caliper/width of 0.25 standard deviations. Exact matching was applied for sex and genotyping array. A total of 1,685 case-control studies with >50 cases were used for our analyses of the MGI phenome.

#### MGI Common Chronic Conditions

We used the CCW Condition Algorithms (rev. 02/2021) from the CMS Chronic Condition Warehouse (CCW; see **Web Resources**) to define 27 common chronic conditions in MGI. In short, like the PheCode system the CCW algorithms are based on ICD-9-CM- and ICD-10-CM-based inclusion and exclusion criteria. Here, we were interested in any observation of such conditions and disregarded the algorithms’ stated reference period or the required numbers/types of qualifying claims for Medicare or Medicaid. The resulting 27 case-control studies were labelled CCW01 – CCW27 and are listed in **Table S12**.

### UK Biobank cohort (UKB)

#### UKB Cohort

UKB is a population-based cohort collected from multiple sites across the United Kingdom and includes over 500,000 participants aged between 40 and 69 years when recruited in 2006–2010 ^101^. The open-access UK Biobank data used in this study included genotypes, ICD9 and ICD10 codes, biomarker data, anthropometrics, vitals, women’s health, health behavior, inferred sex, inferred White British ancestry, kinship estimates down to third degree, birthyear, genotype array, and precomputed principal components of the genotypes. UK Biobank received ethical approval from the NHS National Research Ethics Service North West (11/NW/0382).

#### UKB Genotype Data

We used the UK Biobank Imputed Dataset (v3) and limited analyses to the documented 408,595 White British ^102^ individuals and 47,836,001 variants with imputation information score >= 0.3 and MAF ≥ 0.01% of which 22,933,317 overlapped with the imputed MGI data (see above). Two random subsets of 5,000 and 10,000 unrelated, White British individuals were used for LD analyses of UKB-based summary statistics. Genotyping, quality control, and imputation are described in detail elsewhere ^98^.

#### UKB Phenome

The UK Biobank phenome was based on ICD9 and ICD10 code data of 408,595 White British ^102^, genotyped individuals that were similarly aggregated to PheCodes as MGI (see above, also described elsewhere ^103^). In contrast to MGI, there were many pairwise relationships reported for UKB participants.

To retain a larger effective sample size for each phenotype, we first selected a maximal set of unrelated cases for each phenotype (defined as no pairwise relationship of 3^rd^ degree or closer ^96,104^) before selecting a maximal set of unrelated controls unrelated to these cases. Similar to MGI, we matched up to 10 controls to each case using the R package “MatchIt” ^100^. Nearest neighbor matching was applied for birthyear (as proxy for age because age at diagnosis was not available to us) and PC1-4 (Mahalanobis-metric matching; matching window caliper/width of 0.25 standard deviations), and exact matching was applied for sex and genotyping array. A total of 1,419 matched case-control studies with >50 cases each were used for our analyses of the UK Biobank phenome.

### Exposure data

For a set of 21 continuous and 7 binary exposures for which we could find freely available and complete GWAS summary statistics (see **Exposure GWAS Summary Statistics** below) we extracted the corresponding EHR data as described in **Table S1**. For the binary exposures that are common disorders (type 2 diabetes, hypertension, insomnia, and sleep apnea) we use the PheWAS code-based definitions (see MGI and UKB phenome above; **Table S7**). Survey based measures with multiple responses per person (never/past/current alcohol use and smoking status) were recoded to never / ever responses. Continuous exposures with multiple measurements across multiple visits (e.g., BMI, biomarkers, or blood pressure values) were harmonized as follows: we first removed outliers per individual per trait by using the interquartile range (IQR) criterion. Next, we aggregated multiple entries per individual by using the mean value across multiple entries per person per trait as the value of the corresponding continuous trait. For the UKB cohort, we calculated the estimated Glomerular Filtration Rate (eGFR) on the natural scale using the harmonized serum creatinine values (data field 30700), race and sex information and the Chronic Kidney Disease Epidemiology Collaboration (CKD-EPI) equation ^105^.

### Exposure GWAS Summary Statistics

For each of the 28 exposures, we collected complete GWAS summary statistics from up to four different sources: (1) catalogued GWAS of the NHGRI EBI GWAS Catalog^106^ (2) GWAS from the FinnGen Consortium; (3) published GWAS meta-analyses; and (4) publicly available GWAS summary statistics of phenome x genome screening efforts of the UK Biobank data (Lee and Neale Lab, see **Table S2** and **Web Resources**). We only included GWAS summary statistics of studies that analyzed broad European ancestry to match the ancestry of discovery GWAS and target cohorts (MGI and UKB). If needed, we lifted over coordinates of GWAS summary statistics to human genome assembly GRCh37 (LiftOver, UCSC Genome Browser Store, see **Web Resources**). Entries with missing effect alleles, or effect sizes were excluded. If effect allele frequency (EAF) was reported in the summary statistics, we also compared EAF between the discovery GWAS and the target dataset (MGI and/or UKB). If the proportion of likely flipped alleles (whose RAF deviated more than 15% between the datasets) was above 40% we excluded the GWAS as source for PRS construction. These chosen thresholds were subjective and based on clear differentiation between correct and likely flipped alleles on the two diagonals, as noted frequently in GWAS meta-analyses quality control procedures.

## Statistical Methods

### Heritability Estimation

For each set of GWAS summary statistics from both UK Biobank and non-UK Biobank sources (e.g., FinnGen, GWAS catalog, large meta-analyses), we first estimated the SNP heritability to estimate the proportion of phenotypic variance explained by all measured SNPs based on summary statistics. The estimated SNP heritability represents the upper limit for the prediction performance of PRS methods and serves as an initial filtering criterion to validate the quality of the downloaded summary statistics. To do so, we applied the method MQS (MinQue for Summary statistics), which was implemented in Gemma, to calculate the SNP heritability estimate (see **Web Resources**)^107,108^. MQS estimates the SNP heritability based on the Minimal Norm Quadratic Unbiased Estimation (MINQUE)^109,110^ criterion. Specifically, we first converted the p-values into marginal z-scores, then we used the z-scores as well as 5,000 randomly selected, unrelated samples (reference panel) as input to run Gemma. Finally, we obtained the proportion of variance in phenotypes explained (PVE) estimates from Gemma which corresponds to the SNP heritability estimate. We further filtered out the summary statistics that had negative heritability estimates.

For binary traits with potentially ascertained case-control data, we converted the heritability estimates from the observed scale to the liability scale using the R package “PDRohde/ugnome” and reported population prevalence estimates (**Table S3**) ^111^.

### ExPRS Construction

We constructed the PRS for an individual j in the form PRS*_j_*=∑*_i_β_i_G_ij_* where *i* indexes the included variants for that trait, weight *β_i_* is the log odds ratios retrieved from the external GWAS summary statistics for variant *i,* and *G_ij_* is a continuous version of the measured dosage data for the risk allele of variant *i* in subject *j.* To construct a PRS, one must determine which genetic loci to include in the PRS and their relative weights. We have obtained GWAS summary statistics from several external sources, resulting in several sets of weights for each trait of interest. For each set of weights obtained from GWAS summary statistics from the above-mentioned sources and each trait, we generated for each exposure GWAS up to 5 different PRSs reflecting the 5 implementations of 4 different PRS methods: the C + T (both, best guess genotype [GT] and dosage [DS] version)^25–27^, lassosum^28^, DBSLMM^29^ and PRS-CS^30^ (**Figure 1**).

We summarized the statistical aspects of these construction methods in **Table S22**. The goal of this approach was to compare multiple PRS methods and find the method that works best for the various types of GWAS summary statistics.

#### LD clumping and P-value Thresholding (C+T)

We performed linkage disequilibrium (LD) clumping/pruning of variants with p-values below 0.1 by using the imputed allele dosages of 10,000 randomly selected samples and a pairwise correlation cut-off at r^2^ < 0.1 within 1Mb window. We constructed many different PRS across a fine grid of p-value thresholds. The p-value threshold with the highest pseudo-R^2^ (binary trait) or highest R^2^ (continuous traits) (see **PRS Evaluation** below) was used to define the optimized “Clumping and Thresholding (C & S)” PRS. We applied two approaches for LD clumping: C + T (GT) and C+T (DS). Specifically, the “C+T (GT)” is implemented by plink-1.9 using the best guess genotypes (GT, imputed genotype dosages are rounded to the next integer) for LD calculations, while “C+T (DS)” is implemented in R and considers the uncertainty of imputed genotypes by using the dosage data (DS).

#### Lassosum

Lassosum obtains PRS weights by applying elastic net penalization to GWAS summary statistics and incorporating LD information from a reference panel. Here, we used 5,000 randomly selected, unrelated samples as the LD reference panel. We applied a MAF filter of 1 % and, in contrast to the previous two approaches, only included autosomal variants that overlap between summary statistics, LD reference panel, and target panel. Each “Lassosum” run resulted in up to 76 combinations of the elastic net tuning parameters s and λ, and consequently, in 76 SNP sets with corresponding weights used to construct. We then selected the PRS with the pseudo-R^2^ (binary trait) or highest R^2^ (continuous traits) to define the “Lassosum” PRS (see **PRS Evaluation** below).

#### Deterministic Bayesian Sparse Linear Mixed Model (DBSLMM)

DBSLMM assumes that the true SNP effect sizes derive from a mixture of normal distributions and relies on an efficient deterministic search algorithm for statistical inference. DBSLMM requires both GWAS summary statistics and LD information from a reference panel. Specifically, DBSLMM first selects SNPs with large effect in a deterministic fashion through the C+T procedure and then directly obtains both large SNP effect sizes and small SNP effect sizes through analytic forms. Here, we used 5,000 randomly selected unrelated samples as the LD reference panel. We applied a MAF filter of 1% and only included autosomal variants that overlap between summary statistics, LD reference panel, and target panel. Heritability estimates obtained from Gemma (see above-mentioned procedure) were used as the input of DBSLMM. All other parameters we used are the default parameters in the “DBSLMM” software. For example, we set the cutoff of SNPs clumping and pruning to be r^2^ < 0.1 within a 1Mb window and *p*-value < 1e-06, respectively. Each DBSLMM run resulted in one SNP set with corresponding weights to construct the PRS. We used the default version of DBSLMM which does not require cross-validation and refer to the obtained PRS as “DBSLMM” PRS.

#### PRS-CS

PRS-CS utilizes a Bayesian regression framework and assumes a continuous shrinkage (CS) prior on the effect sizes. Specifically, we applied the default “auto” version of PRS-CS that obtain weights through Gibbs sampling algorithm. Here, PRS-CS-auto uses a precomputed LD reference panel based on external European samples of the 1000 Genomes Project (“EUR reference”) to construct a PRS. We applied a MAF filter of 1 % and only included autosomal variants that overlap between summary statistics, LD reference panel, and target panel. The obtained PRS is referred to as “PRS-CS” PRS.

For each trait and set of GWAS summary statistics, these approaches usually resulted in up to five PRS. However, approaches that resulted in less than 5 weights / variants were excluded. Using the R package “Rprs” (see **Web Resources**), the value of each PRS was then calculated for each MGI participant and, if the GWAS source did to the best of our knowledge did not include UKB samples, also for each UKB participant. For comparability of association effect sizes corresponding to the continuous PRS across exposures and PRS construction methods, we centered PRS values in MGI and UKB to a mean of 0 and scaled them to have a standard deviation of 1.

### ExPRS Evaluation

To assess the predictive performance of these generated PRS, each PRS was assessed through cross-validation in either the MGI cohort or the UKB cohort: we split the data corresponding to each trait in training (50% of the samples with gender ratio unchanged) and test set (50% of the samples with gender ratio unchanged). We used the training set to determine the PRS tuning parameter(s) and used the testing set to obtain performance metric for that PRS.

For the PRS evaluations, except for when computing the pseudo-R^2^ for binary exposures (which is a measure of marginal association of the ExPRS with the exposure)^112^, we fit the following model for each PRS and exposure adjusting for covariates:

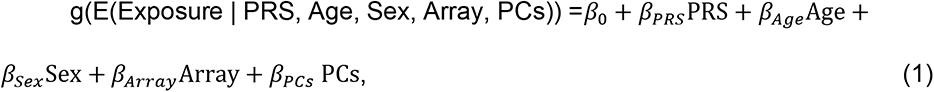

where g(·) is the link function (e.g., identity link function for continuous traits and logit link function for binary traits). PCs were the first four principal components obtained from the principal component analysis of the genotyped GWAS markers, where “Age” was the age at last observed diagnosis in MGI and birthyear in UKB and where “Array” represents the genotyping array.

#### Binary Traits

We used Nagelkerke’s pseudo-R^2^ to select the tuning parameters within the “C+T” and Lassosum construction methods (P-value for “C+T” SNP sets; s and λ for Lassosum) and kept the PRS with the highest pseudo-R^2^ for further analyses. For each PRS derived for each GWAS source/method combination, we assessed the following performance measures relative to observed disease status in MGI and UKB: (1) overall performance with Nagelkerke’s pseudo-R^2^ using R packages “rcompanion”, (2) accuracy with Brier score using R package “DescTools”; and (3) ability to discriminate between cases and controls as measured by the area under the covariate-adjusted receiver operating characteristic (AROC; semiparametric frequentist inference) curve (denoted AAUC) using R package “ROCnReg”. Firth’s bias reduction method was used to resolve the problem of separation in logistic regression (R package “brglm2”).

#### Continuous Traits

For the PRS evaluations of continuous traits, we used R^2^ to select the tuning parameters within the “C+T” and Lassosum construction methods (P-value for “C+T” SNP sets; s and λ for Lassosum) and kept the PRS with the highest R^2^ for further analyses. For each PRS derived for each GWAS source/method combination, we assessed the prediction performance in terms of R^2^ in MGI and UKB.

### ExPRS Primary Association with the Underlying Exposure

Next, we assessed the strength of the relationship between these PRSs and the traits they were designed for. To do this, we fit the same model as equation (1). Our primary interest is *β_PRS_*, while the other factors (Age, Sex and PCs) were included to address potential residual confounding. Firth’s bias reduction method was used to resolve the problem of separation in logistic regression (Logistf in R package “EHR”). As an initial filtering step, we removed PRS that were not significantly associated with their corresponding exposure in MGI or UKB cohort (P > 0.05) for downstream analysis. The majority of these filtered PRS were either based on discovery GWAS with small sample sizes that often did not identify any genome-wide significant hits or were evaluated for exposure with small sample sizes or both, indicating a potential lack of power in our analysis.

### Illustrative Examples showcasing the use of ExPRS

Once we select the ExPRS that were mostly and positively associated with the specific exposure, referred to as the best performing PRS, we use these selected PRSs for various analyses to illustrate how a user may gainfully use these constructs in understanding disease etiology and mechanisms.

#### Phenome-wide Exploration of ExPRS Associations

We conducted PheWAS in MGI and UKB (if the GWAS source was not based on UKB) to identify phenotypes associated with the ExPRS. To evaluate ExPRS-phenotype associations, we conducted Firth bias-corrected logistic regression by fitting the following model for each ExPRS and each phenotype of the corresponding phenome.

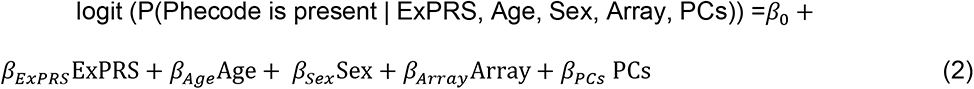

To adjust for multiple testing, we applied the conservative phenome-wide Bonferroni correction according to the total number of analyzed PheCodes (MGI: 1,685 phenotypes; UKB: 1,419 phenotypes as described in **Table S7**). In Manhattan plots, we present –log10 (*p*-value) corresponding to tests for association of the underlying phenotype with the ExPRS. Directional triangles on the PheWAS plot indicate whether a trait was positively (pointing up) or negatively (pointing down) associated with the ExPRS.

To investigate the possibility of the secondary trait associations with ExPRS being completely driven by the exposure or extremes of the trait distribution, we performed a second set of PheWAS: for binary exposures we excluded individuals with the binary exposures for which the ExPRS was constructed; for continuous exposures we excluded individuals with measurements outside of the normal range (**Table S1**). We referred to these PheWAS as “Exclusion-PRS-PheWAS” as described previously^33^.

To evaluate whether the constructed ExPRS is a good proxy for the corresponding exposure, we also repeated the PheWAS using the exposure or normal range exposure as the predictor instead. We referred to these PheWAS as “Trait- PheWAS” and “Exclusion-Trait-PheWAS”, respectively.

#### Utilities of ExPRS on Common Chronic Conditions

To investigate the utility of our constructed ExPRS in predicting Common Chronic Condition in the MGI cohort (see *MGI Common Chronic Conditions* above, **Table S12**). We first split the common chronic conditions into training (50% of the samples with gender ratio unchanged) and test set (50% of the samples with gender ratio unchanged). We conducted Firth bias-corrected logistic regression by fitting the following model for each of the best performing ExPRS and each common chronic condition:

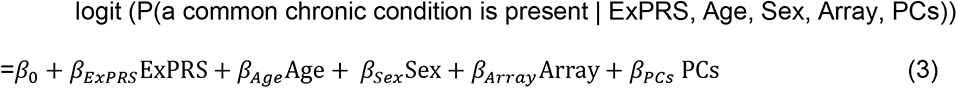

Prediction performance was measured by Nagelkerke’s pseudo-R^2^, Brier score and AAUC. Then we repeated the analysis using the actual exposure as predictor to be trained and evaluated in the MGI cohort.

Next, we selected for each chronic condition the ExPRSs that reached nominal significance in the univariate model and performed clumping (r < 0.5). For each chronic condition, we combined the resulting sets of their associated ExPRSs by fitting a logistic regression in the training set to obtain the linear predictors that we defined as “multiExPRS” in the testing data.

To investigate whether such multiExPRSs can be helpful in predicting a common chronic condition “Y” beyond the condition specific PRS “YPRS” (e.g., breast cancer PRS), we collected the YPRS from public resources, except for type 2 diabetes and hypertension for which we generated ExPRS. We downloaded PRS constructs/weights for lung cancer, prostate cancer, colorectal cancer and breast cancer PRS from Cancer PRSweb^113^ (https://prsweb.sph.umich.edu) and downloaded the following PRS weight from the PGS Catalog^114^ (https://www.pgscatalog.org): Stroke/Transient Ischemic Attach, Heart Failure, Glaucoma, Chronic Kidney Disease, Atrial Fibrillation and Asthma PRS. We harmonized the downloaded PRS weights to GRCh37/hg19 and determined overlap with the MGI genotype data. Non-ambiguous SNP alleles were flipped to the genomic plus strand. We fit three logistic models for each common chronic condition “Y” using the following predictors adjusting for the set of covariates from above: (1) condition specific PRS “YPRS”, (2) the combined ExPRS “multiExPRS”, and (3) “multiExPRS + YPRS”. As before, we combined multiple predictors fitting a logistic regression in the training set to obtain the linear predictors that we used as combined score in the testing data.

To study the ability of these three predictors to identify high risk patients for these chronic conditions, we fit the three multi-variate logistic model but replaced the risk scores with an indicator for whether the risk scores value was in the top 1, 2, 5, 10, or 25% (defined in controls) of the risk score distribution.

#### Poly-Exposure Score Construction and Comparison

To contrast the predictive power of a poly-exposure score (PXS) with combined ExPRSs (multiExPRS, see above), we extracted the collected / measured exposure data from MGI. We removed three exposures (Cystatin C, Fasting Plasma Glucose, Estradiol levels) that due to their high missingness would have led to very small sample sizes in a complete case analysis across multiple exposures.

We retained the training / testing data split from the “ExPRS Evaluation” (see above) and ran the following model for each of the remaining exposures and each of the selected common chronic conditions in the training data:

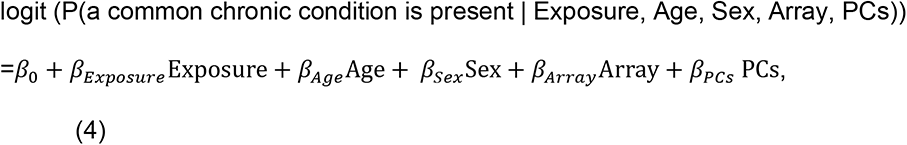

Similar to the multiExPRSs, we selected the significantly associated exposures and performed clumping to only retain the significantly associated exposures with a correlation < 0.5 with each other. The remaining set of exposures was used to create a complete case training data set that we used to obtain effect sizes for each exposure that we used as weights to create weighted exposures in the complete case testing data. The weighted exposures were then combined into a single predictor that we refer to as poly-exposure score (PXS). Finally, we compared the AAUC of following four predictors adjusting for the set of covariates from above: the condition specific PRS (“YPRS”), the combined ExPRSs “multiExPRS”, the “multiExPRS + YPRS” and the PXS.

### Online Visual Catalog: Ex*PRSweb*

The online open access visual catalog *ExPRSweb* (see **Web Resources**) was implemented using Grails as previously described ^20^.

Unless otherwise stated, analyses were performed using R 4.1.1. We used the STREGA checklist when writing our report ^115^.

## Supporting information

STREGA Checklist

## Data Availability

Data cannot be shared publicly due to patient confidentiality. The data underlying the results presented in the study are available from the UK Biobank at http://www.ukbiobank.ac.uk/register-apply/ and from the MGI Study at https://precisionhealth.umich.edu/our-research/michigangenomics/ for researchers who meet the criteria for access to confidential data.

https://exprsweb.sph.umich.edu

## Acknowledgement

The authors acknowledge the Michigan Genomics Initiative participants, Precision Health at the University of Michigan, and the University of Michigan Medical School Data Office for Clinical and Translational Research, the University of Michigan Medical School Central Biorepository, and the University of Michigan Advanced Genomics Core for providing data storage, management, processing, and distribution services, and the Center for Statistical Genetics in the Department of Biostatistics at the School of Public Health for genotype data curation, imputation, and management in support of the research reported in this publication.

We would like to thank Alison Mondul and Brett Vanderwerff (University of Michigan School of Public Health) for careful reading of the manuscript.

Part of this research has been conducted using both the UK Biobank Resource under application number 24460 and using results and data generated by previous researchers who have used the UK Biobank Resource.

This material is based in part upon work supported by the National Institutes of Health/NIH (NCI P30CA046592 [LGF, BM]), by the University of Michigan (UM- Precision Health Investigators Award U063790 [LGF, SP, YM, BM]), and by the National Science Foundation under grant number DMS-1712933. Any opinions, findings, and conclusions or recommendations expressed in this material are those of the author(s) and do not necessarily reflect the views of the National Science Foundation.

## Web Resources

The Michigan Genomics Initiative (MGI), https://precisionhealth.umich.edu/our-research/michigangenomics/

CMS Chronic Condition Warehouse, https://www2.ccwdata.org/web/guest/home/

UCSC Genome Browser Store, https://genome-store.ucsc.edu

Locuszoom, https://github.com/statgen/locuszoom

Rprs, https://github.com/statgen/Rprs

ExPRSweb, https://exprsweb.sph.umich.edu

NHGRI-EBI GWAS Catalog, https://www.ebi.ac.uk/gwas/summary-statistics

FinnGen consortium, https://www.finngen.fi/en/access_results

UKB GWAS (Lee Lab): https://www.leelabsg.org/resources

UKB GWAS (Neale Lab): https://github.com/Nealelab/UK_Biobank_GWAS

Gemma and DBSLMM: https://zlab.org/software.html

lassosum: https://github.com/tshmak/lassosum

PRS-CS: https://github.com/getian107/PRScs

PLINK: https://www.cog-genomics.org/plink2

## References

1. Buniello, A., et al. The NHGRI-EBI GWAS Catalog of published genome-wide association studies, targeted arrays and summary statistics 2019. Nucleic Acids Res 47, D1005–D1012 (2019).

2. Genin, E. Missing heritability of complex diseases: case solved? Hum Genet 139, 103–113 (2020).

3. Manolio, T.A., et al. Finding the missing heritability of complex diseases. Nature 461, 747–53 (2009).

4. Yang, J., et al. Common SNPs explain a large proportion of the heritability for human height. Nature genetics 42, 565–569 (2010).

5. Kamps, R., et al. Next-Generation Sequencing in Oncology: Genetic Diagnosis, Risk Prediction and Cancer Classification. Int J Mol Sci 18(2017).

6. Jostins, L. & Barrett, J.C. Genetic risk prediction in complex disease. Hum Mol Genet 20, R182–8 (2011).

7. Ma, Y. & Zhou, X. Genetic prediction of complex traits with polygenic scores: a statistical review. Trends Genet 37, 995–1011 (2021).

8. Meigs, J.B., et al. Body mass index, metabolic syndrome, and risk of type 2 diabetes or cardiovascular disease. J Clin Endocrinol Metab 91, 2906–12 (2006).

9. Almirall, J., Serra-Prat, M., Bolíbar, I. & Balasso, V. Risk factors for community- acquired pneumonia in adults: a systematic review of observational studies. Respiration 94, 299–311 (2017).

10. Pierce, B.L., Kraft, P. & Zhang, C. Mendelian randomization studies of cancer risk: a literature review. Curr Epidemiol Rep 5, 184–196 (2018).

11. Kachuri, L., et al. Pan-cancer analysis demonstrates that integrating polygenic risk scores with modifiable risk factors improves risk prediction. Nat Commun 11, 6084 (2020).

12. Haneuse, S. Distinguishing Selection Bias and Confounding Bias in Comparative Effectiveness Research. Med Care 54, e23–9 (2016).

13. Beesley, L.J. & Mukherjee, B. Statistical inference for association studies using electronic health records: handling both selection bias and outcome misclassification. Biometrics (2020).

14. Loos, R.J.F. 15 years of genome-wide association studies and no signs of slowing down. Nat Commun 11, 5900 (2020).

15. Liu, M., et al. Association studies of up to 1.2 million individuals yield new insights into the genetic etiology of tobacco and alcohol use. Nat Genet 51, 237–244 (2019).

16. Khera, A.V., et al. Genome-wide polygenic scores for common diseases identify individuals with risk equivalent to monogenic mutations. Nat Genet 50, 1219–1224 (2018).

17. Lambert, S.A., Abraham, G. & Inouye, M. Towards clinical utility of polygenic risk scores. Hum Mol Genet 28, R133–R142 (2019).

18. Tam, C.H.T., et al. Development of genome-wide polygenic risk scores for lipid traits and clinical applications for dyslipidemia, subclinical atherosclerosis, and diabetes cardiovascular complications among East Asians. Genome Med 13, 29 (2021).

19. Ma, Y. & Zhou, X. Genetic prediction of complex traits with polygenic scores: a statistical review. Trends Genet (2021).

20. Fritsche, L.G., et al. Cancer PRSweb: An Online Repository with Polygenic Risk Scores for Major Cancer Traits and Their Evaluation in Two Independent Biobanks. Am J Hum Genet 107, 815–836 (2020).

21. Andrews, S.J., et al. Causal Associations Between Modifiable Risk Factors and the Alzheimer’s Phenome. Ann Neurol 89, 54–65 (2021).

22. Li, S. & Schooling, C.M. A phenome-wide association study of genetically mimicked statins. BMC Med 19, 151 (2021).

23. Richardson, T.G., Harrison, S., Hemani, G. & Davey Smith, G. An atlas of polygenic risk score associations to highlight putative causal relationships across the human phenome. Elife 8(2019).

24. Lambert, S.A., et al. The Polygenic Score Catalog as an open database for reproducibility and systematic evaluation. Nat Genet 53, 420–425 (2021).

25. Wray, N.R., Goddard, M.E. & Visscher, P.M. Prediction of individual genetic risk to disease from genome-wide association studies. Genome Res 17, 1520–8 (2007).

26. International Schizophrenia, C., et al. Common polygenic variation contributes to risk of schizophrenia and bipolar disorder. Nature 460, 748–52 (2009).

27. Euesden, J., Lewis, C.M. & O’Reilly, P.F. PRSice: Polygenic Risk Score software. Bioinformatics 31, 1466–8 (2015).

28. Mak, T.S.H., Porsch, R.M., Choi, S.W., Zhou, X. & Sham, P.C. Polygenic scores via penalized regression on summary statistics. Genet Epidemiol 41, 469–480 (2017).

29. Yang, S. & Zhou, X. Accurate and Scalable Construction of Polygenic Scores in Large Biobank Data Sets. Am J Hum Genet 106, 679–693 (2020).

30. Ge, T., Chen, C.Y., Ni, Y., Feng, Y.A. & Smoller, J.W. Polygenic prediction via Bayesian regression and continuous shrinkage priors. Nat Commun 10, 1776 (2019).

31. Sudlow, C., et al. UK biobank: an open access resource for identifying the causes of a wide range of complex diseases of middle and old age. PLoS medicine 12, e1001779 (2015).

32. Michigan Genomics Initiative (MGI). (2020).

33. Fritsche, L.G., et al. Association of polygenic risk scores for multiple cancers in a phenome-wide study: results from the Michigan Genomics Initiative. The American Journal of Human Genetics 102, 1048–1061 (2018).

34. Visscher, P.M., Hill, W.G. & Wray, N.R. Heritability in the genomics era—concepts and misconceptions. Nature reviews genetics 9, 255–266 (2008).

35. Yang, J., et al. Genetic variance estimation with imputed variants finds negligible missing heritability for human height and body mass index. Nature genetics 47, 1114–1120 (2015).

36. Silventoinen, K., et al. Heritability of adult body height: a comparative study of twin cohorts in eight countries. Twin Research and Human Genetics 6, 399–408 (2003).

37. Johnson, D.A., Billings, M.E. & Hale, L. Environmental determinants of insufficient sleep and sleep disorders: implications for population health. Current epidemiology reports 5, 61–69 (2018).

38. Ge, T., Chen, C.-Y., Ni, Y., Feng, Y.-C.A. & Smoller, J.W. Polygenic prediction via Bayesian regression and continuous shrinkage priors. Nature communications 10, 1–10 (2019).

39. Vilhjálmsson, B.J., et al. Modeling linkage disequilibrium increases accuracy of polygenic risk scores. The american journal of human genetics 97, 576–592 (2015).

40. Kulm, S., Mezey, J. & Elemento, O. Benchmarking the accuracy of polygenic risk scores and their generative methods. MedRxiv 10, 06.20055574 (2020).

41. Pain, O., et al. Evaluation of polygenic prediction methodology within a reference-standardized framework. PLoS genetics 17, e1009021 (2021).

42. Ni, G., et al. A comparison of ten polygenic score methods for psychiatric disorders applied across multiple cohorts. Biological Psychiatry (2021).

43. Choi, S.W. & O’Reilly, P.F. PRSice-2: Polygenic Risk Score software for biobank-scale data. Gigascience 8(2019).

44. Chatterjee, N., Shi, J. & Garcia-Closas, M. Developing and evaluating polygenic risk prediction models for stratified disease prevention. Nat Rev Genet 17, 392–406 (2016).

45. Xue, A., et al. Genome-wide association analyses identify 143 risk variants and putative regulatory mechanisms for type 2 diabetes. Nature communications 9, 1–14 (2018).

46. Liu, M., et al. Association studies of up to 1.2 million individuals yield new insights into the genetic etiology of tobacco and alcohol use. Nature genetics 51, 237–244 (2019).

47. Schillaci, G. & Pucci, G. The dynamic relationship between systolic and diastolic blood pressure: yet another marker of vascular aging? Hypertension research 33, 659–661 (2010).

48. Gavish, B., Ben-Dov, I.Z. & Bursztyn, M. Linear relationship between systolic and diastolic blood pressure monitored over 24 h: assessment and correlates. Journal of hypertension 26, 199–209 (2008).

49. Schillaci, G. & Pucci, G. The dynamic relationship between systolic and diastolic blood pressure: yet another marker of vascular aging? Hypertens Res 33, 659–61 (2010).

50. Tam, C.H.T., et al. Development of genome-wide polygenic risk scores for lipid traits and clinical applications for dyslipidemia, subclinical atherosclerosis, and diabetes cardiovascular complications among East Asians. Genome medicine 13, 1–18 (2021).

51. Timpson, N.J., et al. C-reactive protein levels and body mass index: elucidating direction of causation through reciprocal Mendelian randomization. International journal of obesity 35, 300–308 (2011).

52. Unger, G., Benozzi, S.F., Perruzza, F. & Pennacchiotti, G.L. Triglycerides and glucose index: a useful indicator of insulin resistance. Endocrinología y Nutrición (English Edition) 61, 533–540 (2014).

53. Beesley, L.J., Fritsche, L.G. & Mukherjee, B. An analytic framework for exploring sampling and observation process biases in genome and phenome-wide association studies using electronic health records. Stat Med 39, 1965–1979 (2020).

54. Farmer, R., et al. Promises and pitfalls of electronic health record analysis. Diabetologia 61, 1241–1248 (2018).

55. Gray, N., Picone, G., Sloan, F. & Yashkin, A. The relationship between BMI and onset of diabetes mellitus and its complications. Southern medical journal 108, 29 (2015).

56. Wolk, R., Shamsuzzaman, A.S. & Somers, V.K. Obesity, sleep apnea, and hypertension. Hypertension 42, 1067–1074 (2003).

57. Wolfe, B.M., Kvach, E. & Eckel, R.H. Treatment of obesity: weight loss and bariatric surgery. Circulation research 118, 1844–1855 (2016).

58. Shivakumar, S., Srivastava, A. & G, C.S. Body Mass Index and Dental Caries: A Systematic Review. Int J Clin Pediatr Dent 11, 228–232 (2018).

59. Coutinho, T., et al. Central obesity and survival in subjects with coronary artery disease: a systematic review of the literature and collaborative analysis with individual subject data. J Am Coll Cardiol 57, 1877–86 (2011).

60. Ng, R., Sutradhar, R., Yao, Z., Wodchis, W.P. & Rosella, L.C. Smoking, drinking, diet and physical activity—modifiable lifestyle risk factors and their associations with age to first chronic disease. International journal of epidemiology 49, 113–130 (2020).

61. Wynder, E.L., et al. Screening for risk factors for chronic disease in children from fifteen countries. Preventive medicine 10, 121–132 (1981).

62. Chronic Conditions Data Warehouse. CCW Chronic Condition Categories. Vol. 2021.

63. Xu, W., et al. Meta-analysis of modifiable risk factors for Alzheimer’s disease. J Neurol Neurosurg Psychiatry 86, 1299–306 (2015).

64. Choi, S.W. & O’Reilly, P.F. PRSice-2: Polygenic Risk Score software for biobank-scale data. Gigascience 8, giz082 (2019).

65. He, Y., et al. Comparisons of Polyexposure, Polygenic, and Clinical Risk Scores in Risk Prediction of Type 2 Diabetes. Diabetes Care 44, 935–943 (2021).

66. Caldwell, M., Martinez, L., Foster, J.G., Sherling, D. & Hennekens, C.H. Prospects for the Primary Prevention of Myocardial Infarction and Stroke. J Cardiovasc Pharmacol Ther 24, 207–214 (2019).

67. Reis, J.P., et al. Lifestyle factors and risk for new-onset diabetes: a population-based cohort study. Ann Intern Med 155, 292–9 (2011).

68. Guilbert, J.J. The world health report 2002 - reducing risks, promoting healthy life. Educ Health (Abingdon) 16, 230 (2003).

69. Ellingjord-Dale, M., et al. Coffee consumption and risk of breast cancer: A Mendelian randomization study. Plos one 16, e0236904 (2021).

70. Grosso, G., et al. Coffee consumption and risk of all-cause, cardiovascular, and cancer mortality in smokers and non-smokers: a dose-response meta-analysis. (Springer, 2016).

71. Xu, J., et al. Leukocyte telomere length is associated with aggressive prostate cancer in localized prostate cancer patients. EBioMedicine 52, 102616 (2020).

72. Soleimani, A., et al. Role of the transforming growth factor-β signaling pathway in the pathogenesis of colorectal cancer. Journal of cellular biochemistry 120, 8899–8907 (2019).

73. Kubiczkova, L., Sedlarikova, L., Hajek, R. & Sevcikova, S. TGF-β–an excellent servant but a bad master. Journal of translational medicine 10, 1–24 (2012).

74. Wang, H., Peng, R., Wang, J., Qin, Z. & Xue, L. Circulating microRNAs as potential cancer biomarkers: the advantage and disadvantage. Clinical epigenetics 10, 1–10 (2018).

75. Lloyd-Jones, L.R., et al. Improved polygenic prediction by Bayesian multiple regression on summary statistics. Nat Commun 10, 5086 (2019).

76. Prive, F., Arbel, J. & Vilhjalmsson, B.J. LDpred2: better, faster, stronger. Bioinformatics (2020).

77. Chun, S., et al. Non-parametric Polygenic Risk Prediction via Partitioned GWAS Summary Statistics. Am J Hum Genet 107, 46–59 (2020).

78. Prive, F., Vilhjalmsson, B.J., Aschard, H. & Blum, M.G.B. Making the Most of Clumping and Thresholding for Polygenic Scores. Am J Hum Genet 105, 1213–1221 (2019).

79. Marquez-Luna, C., et al. LDpred-funct: incorporating functional priors improves polygenic prediction accuracy in UK Biobank and 23andMe data sets. bioRxiv DOI: https://doi.org/10.1101/375337, 375337 (2020).

80. Hu, Y., et al. Leveraging functional annotations in genetic risk prediction for human complex diseases. PLoS Comput Biol 13, e1005589 (2017).

81. Maier, R., et al. Joint analysis of psychiatric disorders increases accuracy of risk prediction for schizophrenia, bipolar disorder, and major depressive disorder. Am J Hum Genet 96, 283–94 (2015).

82. Martin, A.R., et al. Human Demographic History Impacts Genetic Risk Prediction across Diverse Populations. Am J Hum Genet 100, 635–649 (2017).

83. Martin, A.R., et al. Clinical use of current polygenic risk scores may exacerbate health disparities. Nat Genet 51, 584–591 (2019).

84. Fritsche, L.G., et al. On Cross-ancestry Cancer Polygenic Risk Scores. medRxiv (2021).

85. Sirugo, G., Williams, S.M. & Tishkoff, S.A. The Missing Diversity in Human Genetic Studies. Cell 177, 26–31 (2019).

86. Fritsche, L.G., et al. On Cross-ancestry Cancer Polygenic Risk Scores. medRxiv, 2021.02.24.21252351 (2021).

87. Zeng, P., Shao, Z. & Zhou, X. Statistical methods for mediation analysis in the era of high-throughput genomics: Current successes and future challenges. Comput Struct Biotechnol J 19, 3209–3224 (2021).

88. Guo, Y., et al. Genetically predicted body mass index and breast cancer risk: Mendelian randomization analyses of data from 145,000 women of European descent. PLoS medicine 13, e1002105 (2016).

89. Pierce, B.L., Kraft, P. & Zhang, C. Mendelian randomization studies of cancer risk: a literature review. Current epidemiology reports 5, 184–196 (2018).

90. Shen, X., et al. A phenome-wide association and Mendelian Randomisation study of polygenic risk for depression in UK Biobank. Nat Commun 11, 2301 (2020).

91. Richardson, T.G., Harrison, S., Hemani, G. & Davey Smith, G. An atlas of polygenic risk score associations to highlight putative causal relationships across the human phenome. Elife 8, e43657 (2019).

92. Beesley, L.J., et al. The emerging landscape of health research based on biobanks linked to electronic health records: Existing resources, statistical challenges, and potential opportunities. Stat Med 39, 773–800 (2020).

93. Wang, C., et al. Ancestry estimation and control of population stratification for sequence-based association studies. Nat Genet 46, 409–15 (2014).

94. Li, J.Z., et al. Worldwide human relationships inferred from genome-wide patterns of variation. Science 319, 1100–4 (2008).

95. Manichaikul, A., et al. Robust relationship inference in genome-wide association studies. Bioinformatics 26, 2867–73 (2010).

96. Abraham, K.J. & Diaz, C. Identifying large sets of unrelated individuals and unrelated markers. Source Code Biol Med 9, 6 (2014).

97. McCarthy, S., et al. A reference panel of 64,976 haplotypes for genotype imputation. Nat Genet 48, 1279–83 (2016).

98. Fritsche, L.G., et al. Association of Polygenic Risk Scores for Multiple Cancers in a Phenome-wide Study: Results from The Michigan Genomics Initiative. Am J Hum Genet 102, 1048–1061 (2018).

99. Carroll, R.J., Bastarache, L. & Denny, J.C. R PheWAS: data analysis and plotting tools for phenome-wide association studies in the R environment. Bioinformatics 30, 2375–6 (2014).

100. Ho, D.E., Imai, K., King, G. & Stuart, E.A. MatchIt: Nonparametric Preprocessing for Parametric Causal Inference. Journal of Statistical Software 42, 1–28 (2011).

101. Sudlow, C., et al. UK biobank: an open access resource for identifying the causes of a wide range of complex diseases of middle and old age. PLoS Med 12, e1001779 (2015).

102. Bycroft, C., et al. Genome-wide genetic data on ∼500,000 UK Biobank participants. bioRxiv (2017).

103. Zhou, W., et al. Efficiently controlling for case-control imbalance and sample relatedness in large-scale genetic association studies. Nat Genet 50, 1335–1341 (2018).

104. Michailidou, K., et al. Association analysis identifies 65 new breast cancer risk loci. Nature 551, 92–94 (2017).

105. Levey, A.S., et al. A new equation to estimate glomerular filtration rate. Ann Intern Med 150, 604–12 (2009).

106. Buniello, A., et al. The NHGRI-EBI GWAS Catalog of published genome-wide association studies, targeted arrays and summary statistics 2019. Nucleic acids research 47, D1005–D1012 (2019).

107. Zhou, X. A unified framework for variance component estimation with summary statistics in genome-wide association studies. The annals of applied statistics 11, 2027 (2017).

108. Zhou, X. & Stephens, M. Genome-wide efficient mixed-model analysis for association studies. Nature genetics 44, 821–824 (2012).

109. Rao, C.R. Estimation of variance and covariance components—MINQUE theory. Journal of multivariate analysis 1, 257–275 (1971).

110. Rao, C.R. Estimation of heteroscedastic variances in linear models. Journal of the American Statistical Association 65, 161–172 (1970).

111. Lee, S.H., Wray, N.R., Goddard, M.E. & Visscher, P.M. Estimating missing heritability for disease from genome-wide association studies. Am J Hum Genet 88, 294–305 (2011).

112. Nagelkerke, N.J. A note on a general definition of the coefficient of determination. Biometrika 78, 691–692 (1991).

113. Fritsche, L.G., et al. Cancer PRSweb: an online repository with Polygenic Risk Scores for major cancer traits and their evaluation in two independent biobanks. The American Journal of Human Genetics 107, 815–836 (2020).

114. Lambert, S.A., et al. The Polygenic Score Catalog as an open database for reproducibility and systematic evaluation. Nature Genetics 53, 420–425 (2021).

115. Little, J., et al. STrengthening the REporting of Genetic Association Studies (STREGA): an extension of the STROBE statement. PLoS Med 6, e22 (2009).

