## Supplemental Figures for "ExPRSweb - An Online Repository with Polygenic Risk Scores for Common Health-related Exposures"

**Supplementary Figure 1. Descriptive statistics comparing traits in MGI and UKB.** For continuous traits, the plot of distribution density was shown. For binary traits, box plots were shown.

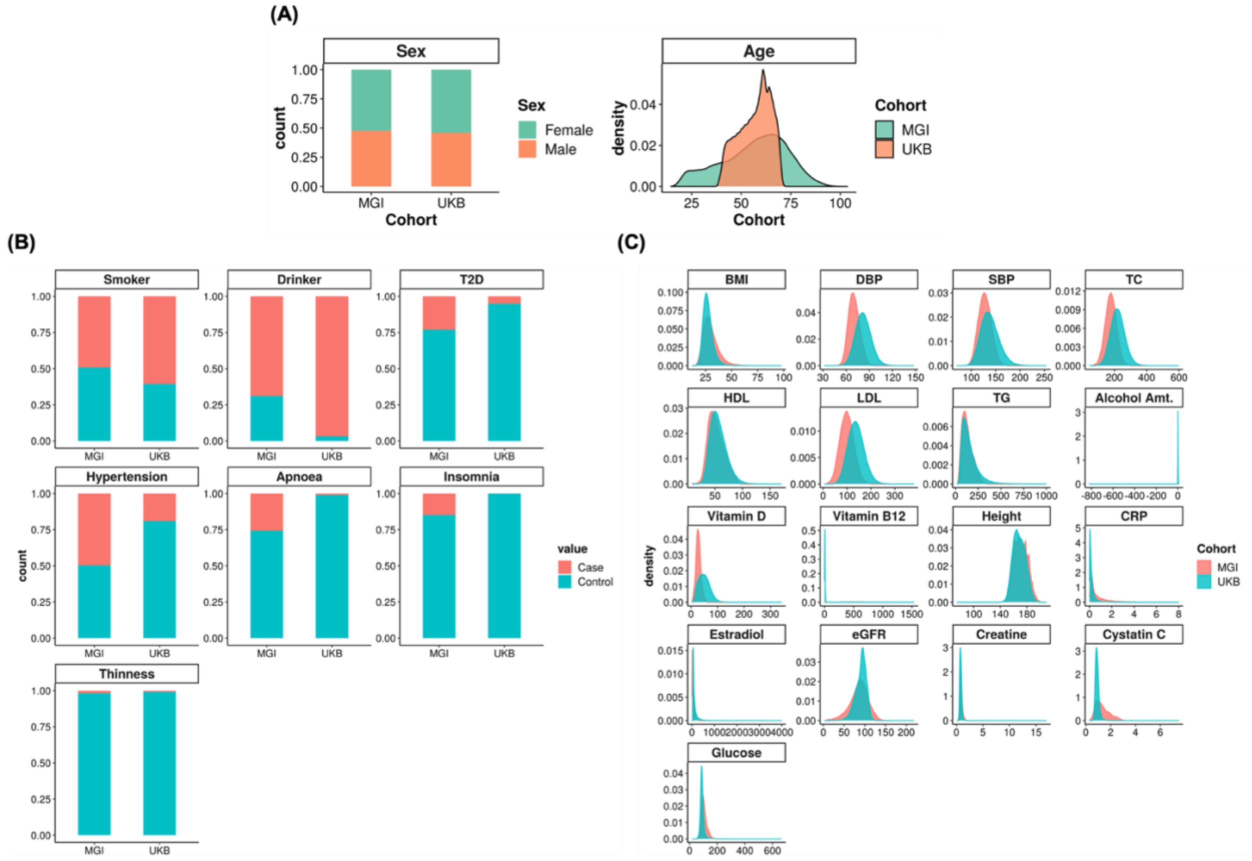

**Supplementary Figure 2. Heritability estimates of 79 collected summary statistics with positive heritability estimates on the liability scale.** Here, three summary statistics were excluded due to the negative estimate on the heritability. Detailed statistics see **Table S3**.

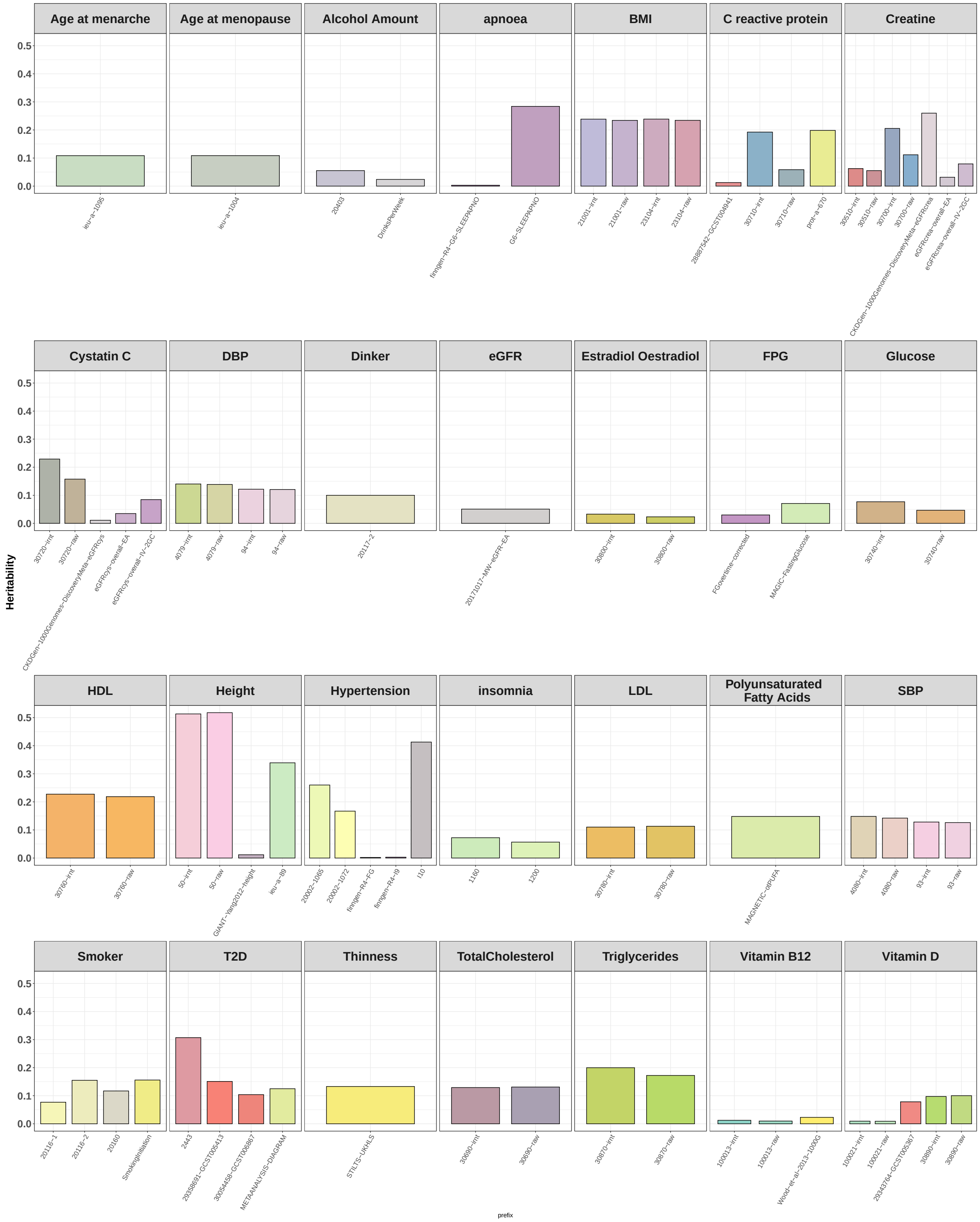

**Supplementary Figure 3. Rank plot of prediction performance of different PRS methods in MGI across ten continuous (left) and six binary traits (right).** For each trait, all methods were ranked according to their prediction performance ( $R^2$ for continuous traits and covariates adjusted AUC for binary traits) in MGI traits. The summed number of achieved ranks is depicted by color and circle size. Method that generated non-significant PRS were ranked as five. For a fair comparison we selected the same summary statistic for each method (GWAS with the highest heritability estimate).

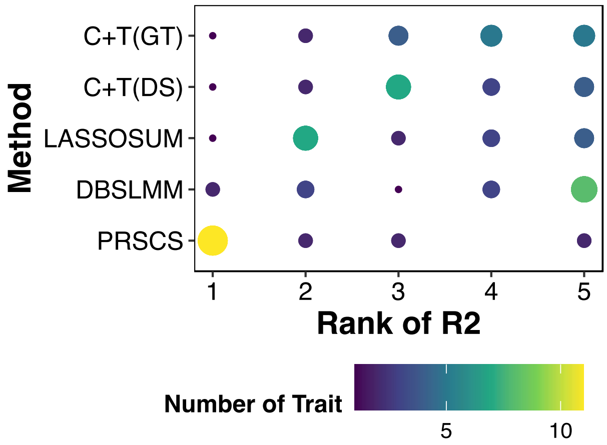


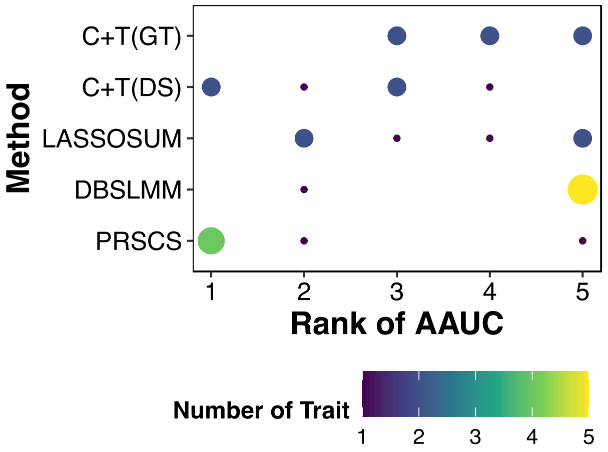

**Supplementary Figure 4. The relationship between heritability estimates and prediction $R^2$ for the summary statistics that generate the best PRS for a specific quantitative exposure trait in MGI cohort.**

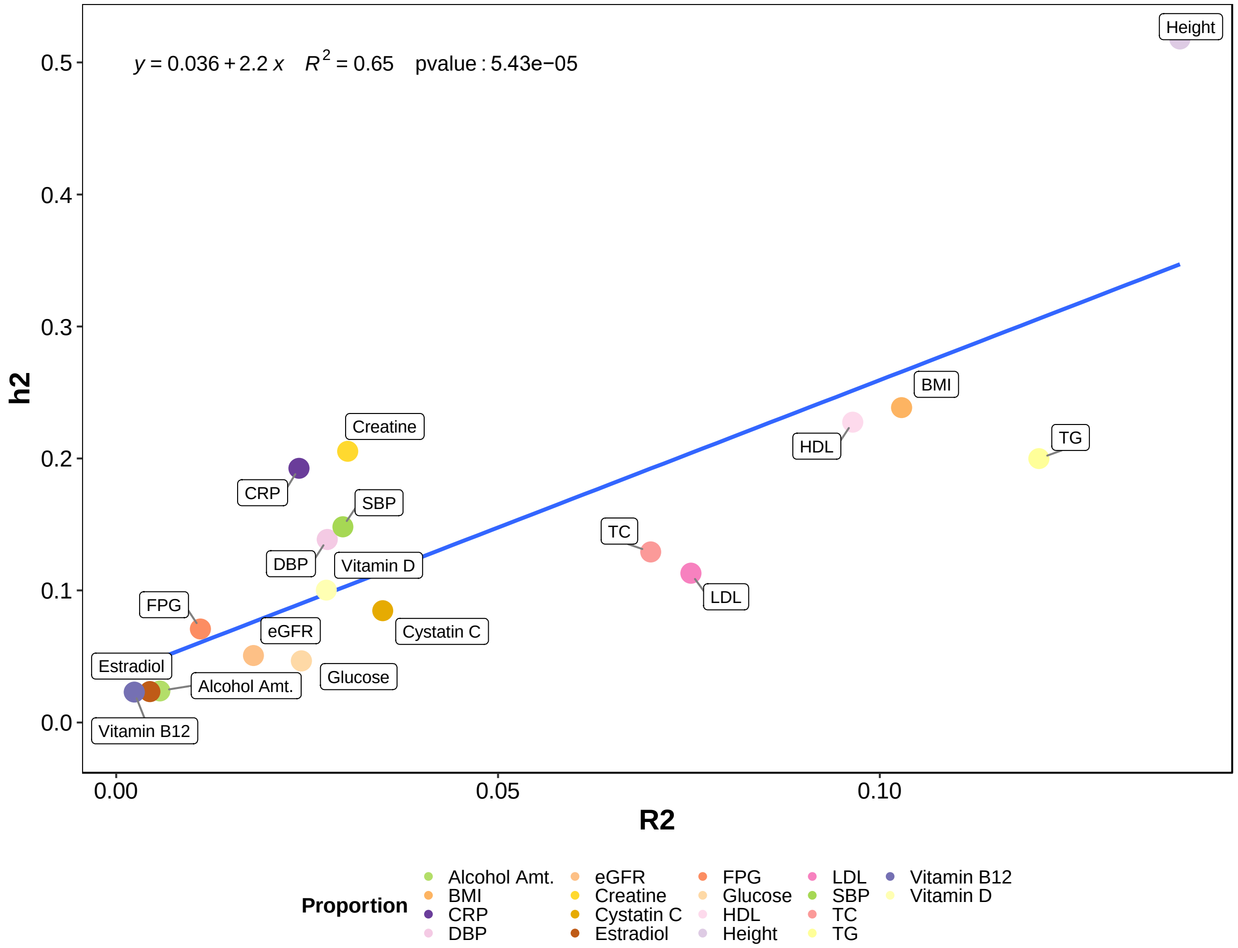

**Supplementary Figure 5. The relationship between the rank of heritability estimates and the rank for prediction $R^2$ of the constructed PRS from each summary statistic across 18 quantitative traits in MGI.**

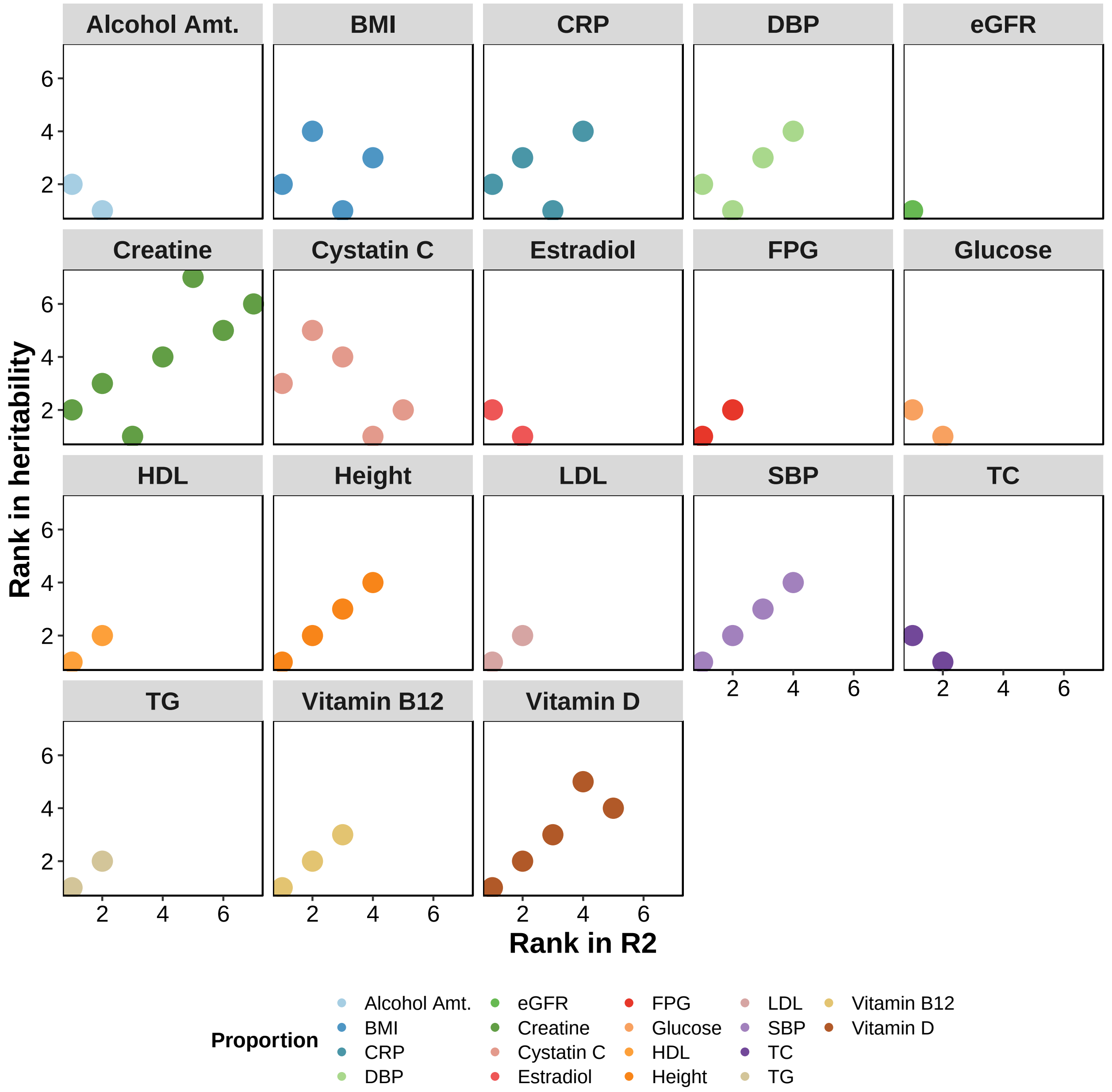

**Supplementary Figure 6. Prediction performance of different PRS methods in UKB across traits.** (A) The color represents the absolute prediction performance for each method across traits. (B) The color is scaled to 0-1 range and represents the relative prediction performance for each method across traits. For both (A) and (B), the prediction performance is quantified as $R^2$ for continuous traits AUC for binary traits. We keep the order of the traits corresponding to the same order of traits in MGI evaluation. For the traits that are not available or the traits that have no best performing PRS in MGI (e.g., age menarche, age menopause, and Cystatin C), we order them by their maximum heritability estimates. Here the summary statistic that generates the best performing exposure PRS was selected for comparison.

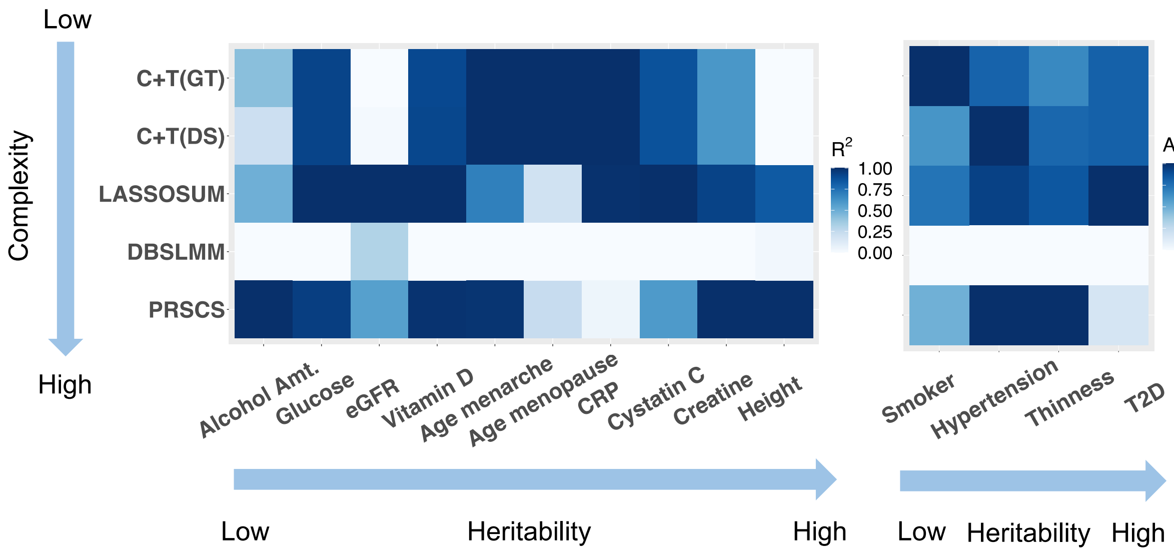

**Supplementary Figure 7. Rank plot of prediction performance of different PRS methods in UKB across traits.** For each trait, all methods were ranked according to their prediction performance ($R^2$ for continuous traits and AUC for binary traits) in UKB traits. The unique ranks each method achieved is shown, colored according to the number of diseases corresponding to that rank. For the method that generated non-significant PRS, we rank it as five. Here the summary statistic that generates the best performing exposure PRS was selected for comparison.

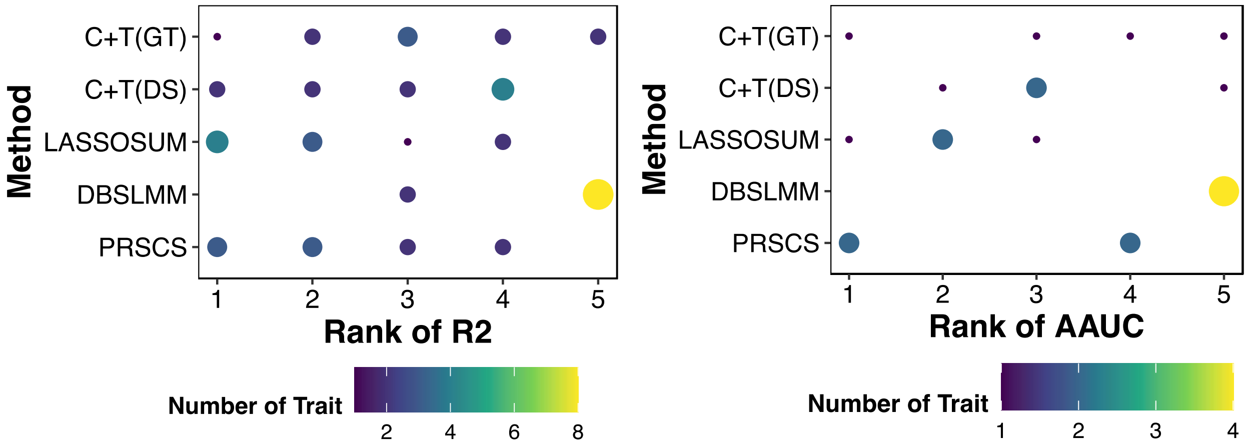

**Supplementary Figure 8. PRS PheWAS and exclusion PRS PheWAS for an example binary trait in MGI.** (A) PRS PheWAS plot is shown for type 2 diabetes PRS predictor (B) Exclusion PRS PheWAS plot is shown for non-type 2 diabetes PRS predictor (C) PRS PheWAS plot is shown for type 2 diabetes predictor (D) Exclusion PRS PheWAS plot is shown for non-type 2 diabetes.

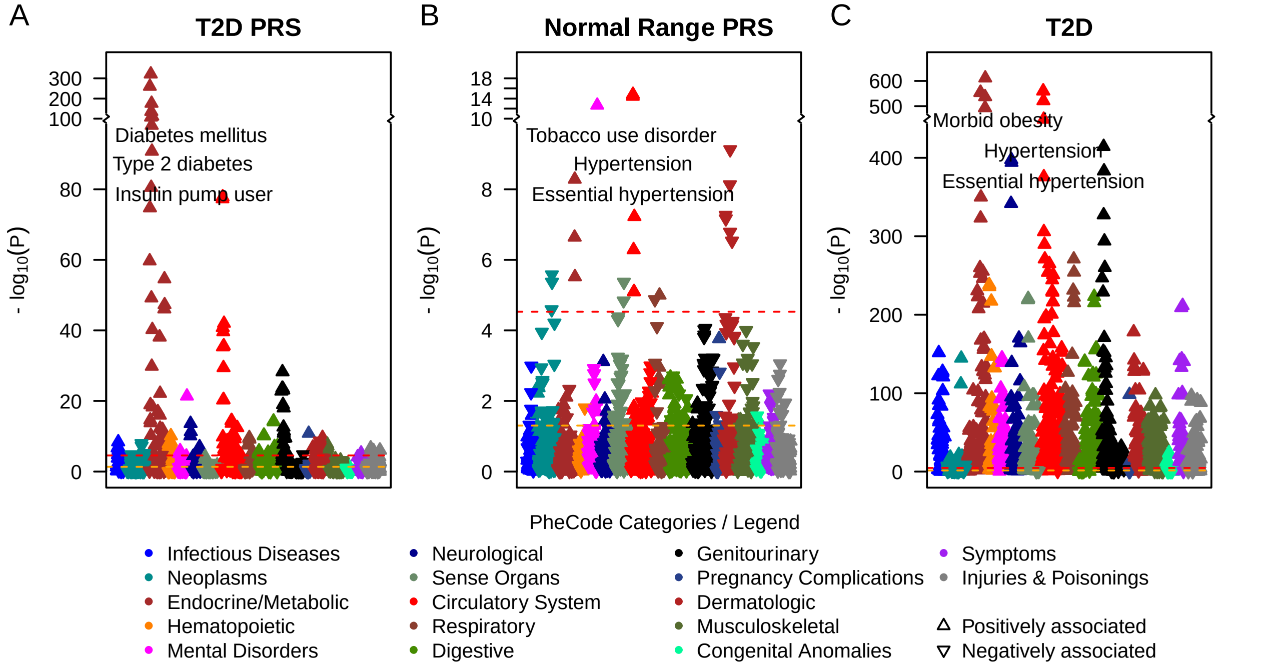

**Supplementary Figure 9. PRS PheWas and exclusion PRS PheWas for example binary trait in UKB.** (A) PRS PheWAS plot is shown for T2D PRS predictor (B) Exclusion PRS PheWAS plot is shown for non- T2D PRS predictor (C) Trait PheWAS plot is shown for T2D predictor

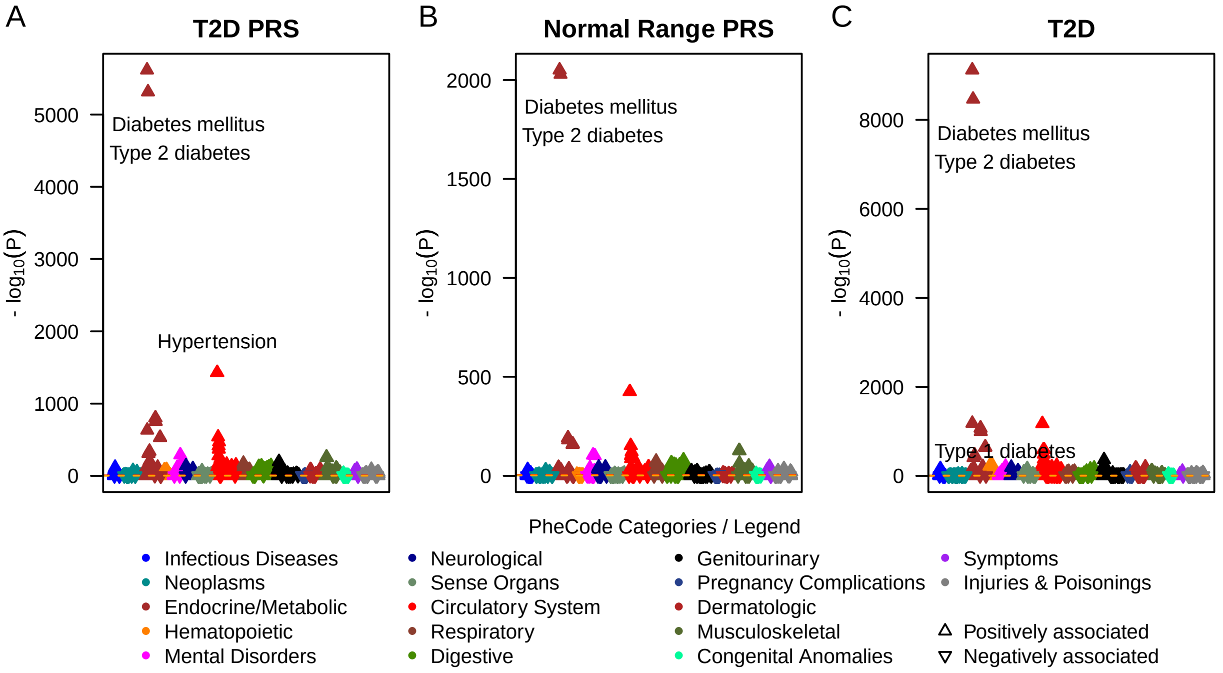

**Supplementary Figure 10. Utility of exposure PRS in risk stratification for 12 chronic conditions.** Exposure PRS can improve the risk stratification compared with using trait specific PRS alone.

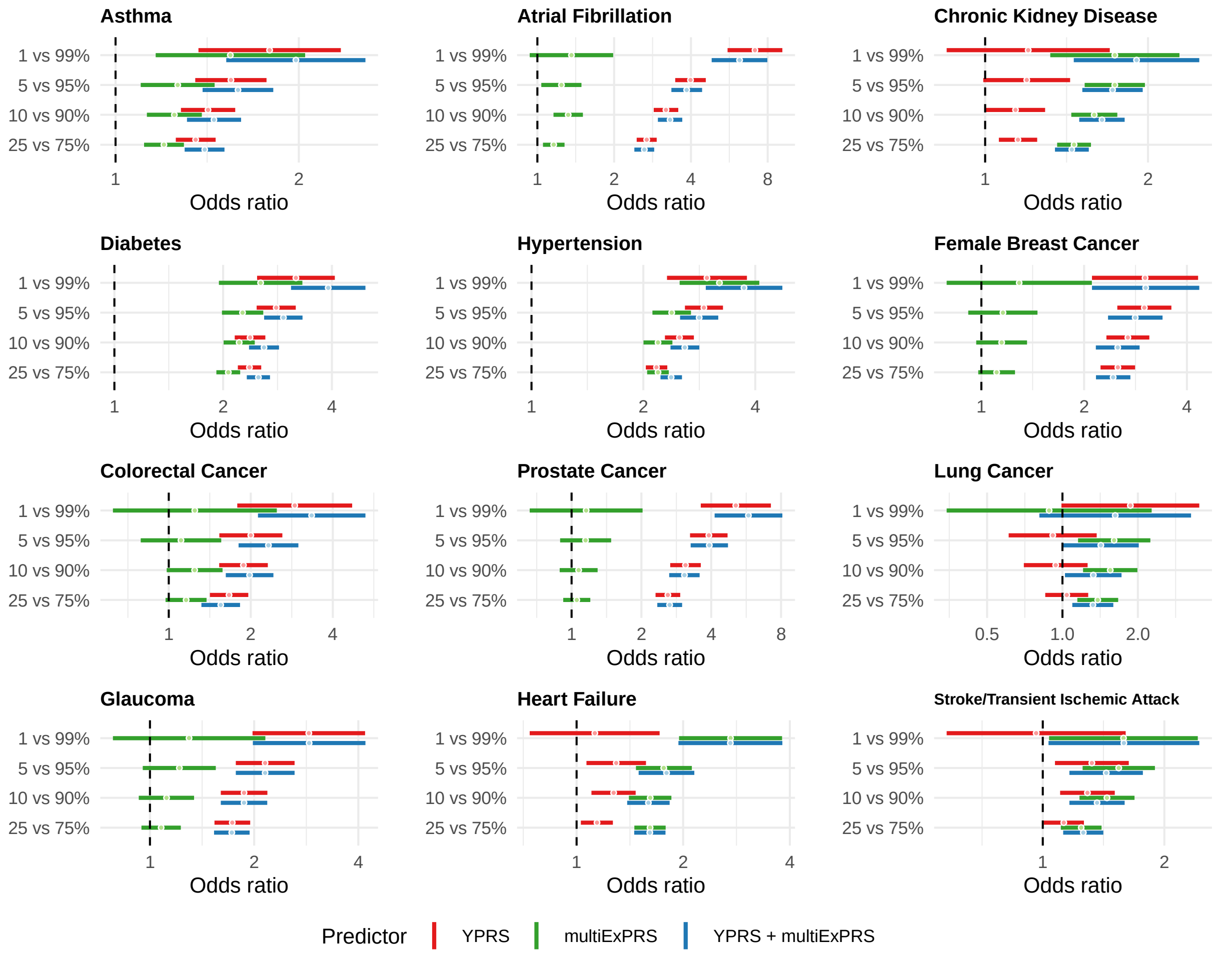

**Supplementary Figure 11. Comparisons of the prediction performance of genetic predictors and the exposure trait score (PXS) for common chronic conditions in MGI cohort.** AAUC paired with 95% confidence interval for condition PXS (green), specific PRS (red), exposure PRS (blue) and trait + exposure PRS (orange) were shown in the format of forest plot. Each bar represents the 95% interval for the AAUC with the dot represents the AAUC estimate.

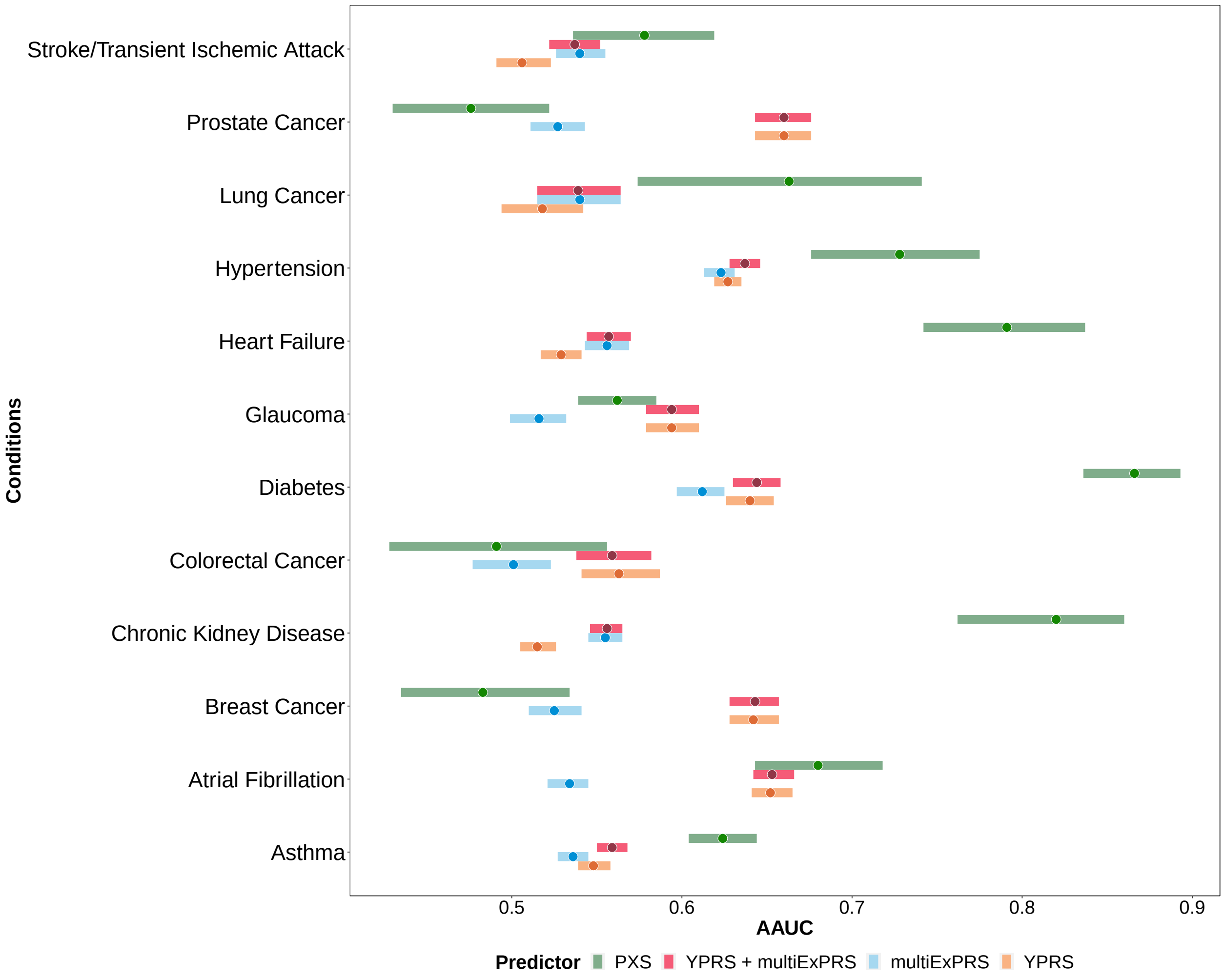
